# Language Modeling Screens Parkinson’s Disease with Self-reported Questionnaires

**DOI:** 10.1101/2024.09.23.24314200

**Authors:** Diego Machado Reyes, Juergen Hahn, Li Shen, Pingkun Yan

**Author notes:** Contributing authors.

## Abstract

Parkinson’s disease (PD) is a growing public health challenge associated with the aging population. Current diagnostic methods rely on motor symptoms and invasive procedures, making early detection difficult. This study established a transferable artificial intelligence (AI) model, Quest2Dx, to analyze health questionnaires to enable low-cost and non-invasive PD diagnosis. Quest2Dx tackles the common challenges of missing responses and required specific modeling for each questionnaire by developing a novel language modeling approach to allow the model transfer across different questionnaires and to enhance the interpretability. Evaluated on the PPMI and Fox Insight datasets, Quest2Dx achieved AUROCs of 0.977 and 0.974, respectively, significantly outperforming existing methods. Additionally, cross-questionnaire validation achieved AUROCs of 0.920 and 0.952, respectively, from PPMI to Fox Insight and vice versa. Quest2Dx also identified key predictors from the list of questions to provide further insights. The validated technology elucidates a promising path for PD screening in primary-care settings.

## 1 Introduction

Parkinson’s disease (PD) is one of the most common neurodegenerative disorders, affecting approximately one million Americans and seven million people worldwide [1, 2]. Given that age is a significant risk factor for PD [2], the prevalence of this disease is expected to rise in tandem with the aging global population, making PD a pressing global public health concern [1]. PD goes often undetected until its intermediate and late stages when characteristic motor symptoms, such as tremors, rigidity, and bradykinesia, become apparent [3]. This delay in diagnosis is partly due to the difficulty of recognizing early stage indicators such as olfactory dysfunction, sleep disturbances, writing changes, and mood changes. These early symptoms may be ignored by patients and primary healthcare givers, or perceived as innocuous when reported separately [4]. This can lead to a lack of timely referrals to expert neurologists [5]. Furthermore, gold-standard methods for diagnosing PD in its early stages, such as cerebrospinal fluid analysis and advanced imaging techniques, are typically invasive and not feasible for population screening [6]. As a result, there is a critical need for developing non-invasive, early diagnostic methods that can be seamlessly adapted to current practices in primary care settings to identify PD before significant motor impairment occurs.

Health questionnaires are routinely administered to patients in primary care settings, offering a valuable opportunity to detect early signs of disease development. By analyzing these responses, patients at risk of conditions like PD could be identified earlier and referred to specialists for further evaluation. These questionnaires are particularly promising for PD screening, as they are non-invasive, cost-effective, require minimal patient effort, and can be easily integrated into standard clinical workflows. However, current reliance on clinicians to interpret health questionnaires can introduce biases, leading to the dismissal of subtle early symptoms due to the overwhelming number of questions and conditions being evaluated simultaneously.

Artificial intelligence (AI) has enormous potential to positively impact primary care practices [7] and could enhance early detection of PD. AI methods, such as machine learning and deep learning approaches, have shown promising results for early PD diagnosis using data from cerebrospinal fluid (CSF) and blood tests [8], genetics [9], wearable technology [10] and brain imaging [11]. However, the use of AI on health questionnaires for PD screening remain largely unexplored.

Health questionnaires come with challenges of their own for AI, as shown in Fig. 1a. They often have missing entries, are difficult to interpret by AI models due to their categorical nature, and have higher levels of noise because of self-assessments. The self-completing nature of questionnaires often leads to partially completed forms. Conventional machine learning methods usually impute the missing entries, which introduces errors in the data [12]. These errors make accurate modeling of the hidden patterns a complex task. At the same time, imputation strategies and machine learning models in general miss to encode the meaning of each question as they only focus on the numerical values from the answers. Additionally, previous methods are not transferable across diverse questionnaires, where questions might be similar but the wording and the answer choices are different. Therefore, there is a crucial need to develop better strategies that can accurately model the missing entries and fully take advantage of the knowledge available through the health questionnaire. Here, large language models present a great opportunity as they can encode the meaning of each question into a numerical representation, that can then be used to provide context to each answer.

**Fig. 1:**
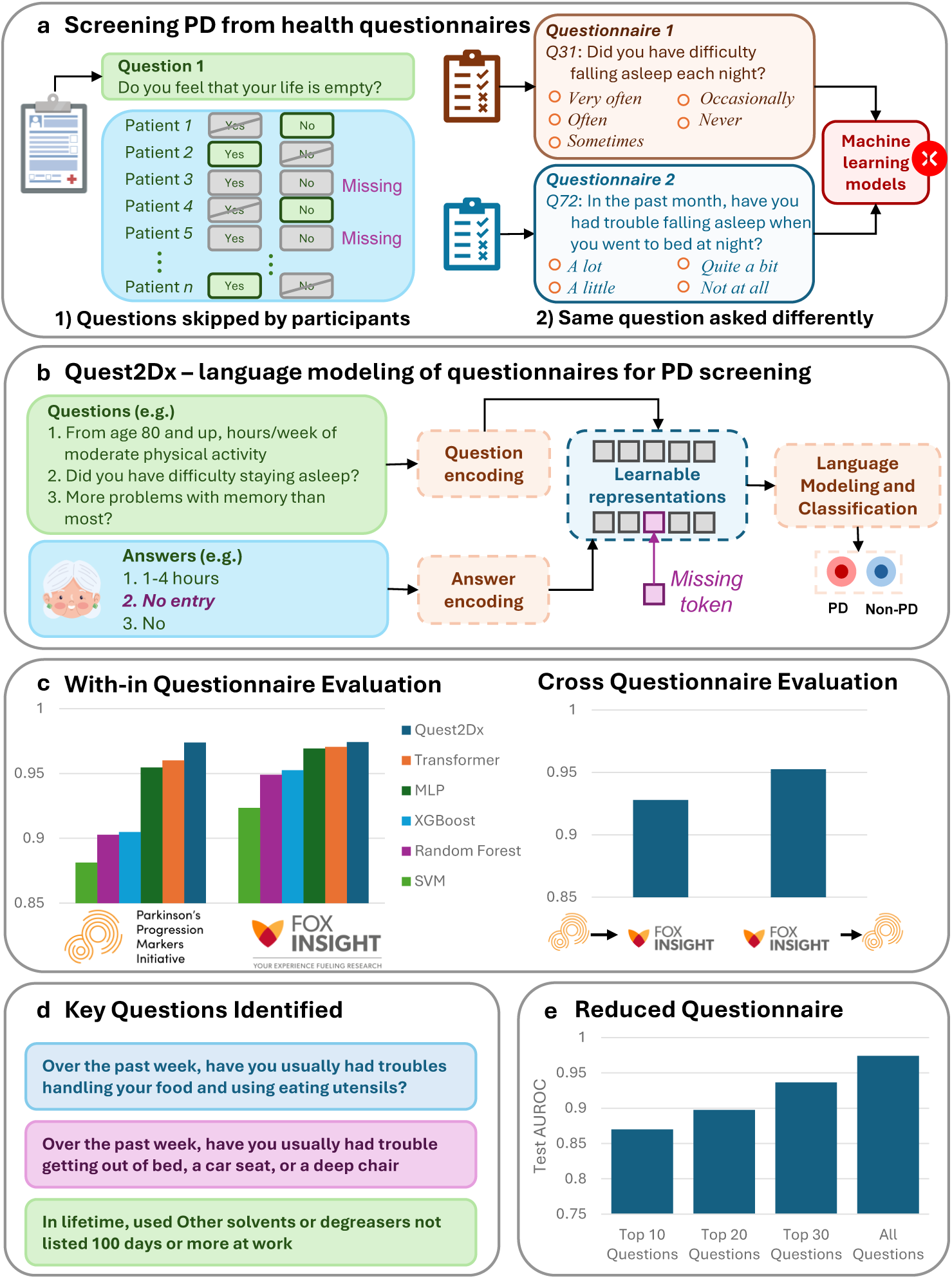
Graphical overview of Quest2Dx. **a.** Self-reported data often contains large volumes of missing entries which makes it very challenging for its analysis. At the same time, health-questionnaires might ask similar questions; however with different wording or options. This limits the applicability of machine learning models across questionnaires. **b.** Quest2Dx uses language modeling to incorporate knowledge from the questions together with the answers given by the patients. It also learns a missing entry data representation to model the missing answers. **c.** Quest2Dx effectively classifies PD patients from healthy controls on two datasets. Quest2Dx is also able to be trained on one questionnaire and be evaluated on a different one. **d.** Quest2Dx is highly interpretable as it is able to identify the questions driving the classification. **e.** A targeted dataset of the top 30 questions identified by Quest2Dx showed strong performances to classify PD patients.

In this work, we developed Quest2Dx, a novel deep learning framework based on the transformer architecture that accurately differentiates PD subjects from healthy controls (HC) using only data collected from health questionnaires. Quest2Dx, as ilustrated in Fig. 1b addresses three key technical challenges and provides interpretable results for PD diagnosis. First, it is able to effectively deal with missing entries and incorporates knowledge from the questions in order to better model the relationships existing between the patients’ answers. Second, its novel design allows Quest2Dx to be trained on one questionnaire and effectively classify patients from a different questionnaire, even when the questions might have been worded differently. Third, Quest2Dx explicitly calculates the most relevant questions-answers pairs for the diagnosis of each subject; thus enhancing its interpretability. Quest2Dx presents two key innovations. First, it introduces a special learnable representation that allows to model the missing entries in the data without preprocessing imputation. Second, it uses a large language model, namely BERT [13], to encode the questions to infuse knowledge to each answer regarding its meaning.

We evaluated all models using two landmark cohorts for PD, the Parkinson’s Progression Markers Initiative (PPMI) Online Study [14] and Fox Insight [15]. PPMI Online is a study from the larger consortium of PPMI, in which the authors have collected participant-reported health information from people with and without PD. The primary goal is to better understand risk and predictive factors for PD. Additionally, they sought to find associations between participant reported outcomes (PROs) provided in PPMI Online with data collected in the PPMI companion studies and the Fox Insight study. PPMI online uses a very extensive collection of questionnaires with a total of 1,387 questions and collected data from 40,012 subjects, 27,802 PD patients and 22,607 healthy controls. Fox Insight is a large online cohort dedicated to study PD patients and their genetic make up. Fox Insight has collected answers online of a similarly extensive set of questionnaires with a total of 7,651 questions from 54,571 subjects, 38,514 with PD and 16,057 healthy controls. Among multiple benchmarking methods, Quest2Dx achieved the best performance with a average area under the receiver operating characteristic curve (AUROC) of 0.977 (95% CI: [0.976,0.978]) for PPMI and 0.974 (95% CI: [0.974, 0.975]) for the Fox Insight dataset, significantly outperforming all alternative methods, as shown in Fig. 1c More importantly, Quest2Dx demonstrated robust transferability, achieving an average AUROC of 0.920 (95% CI: [0.741, 1.115]) when trained on PPMI and evaluated on Fox Insight, and 0.952 (95% CI: [0.925, 0.980]) when trained on Fox Insight and evaluated on PPMI. This strong performance, despite differences in questions between the datasets, underscores the model’s capacity to generalize well across distinct cohorts and questionnaires. The results of Quest2Dx demonstrate that PD can be accurately classified using only self-reported assessments from health questionnaires.

Furthermore, Quest2Dx helped identify the key questions for each questionnaire that enabled the PD patient classification, illustrated in 1d. Prodromal factors such as sleep disorders, and early fine-motor skill loss were found to be good predictors. Similarly, exposure to solvents and degreasers was highly ranked as a risk factor. We demonstrated that a reduced set of the top 30 questions identified by Quest2Dx was able to consistently classify PD patients with an average AUROC of 0.937 (95% CI: [0.935, 0.938]), as shown in 1e. These findings will help design simplified questionnaires for PD screening, paving the way for a tool that can be adopted in future studies and used in primary care settings to enable early diagnosis and timely treatment for patients.

## 2 Results

Quest2Dx and competing benchmarks were trained and evaluated on the questionnaire datasets of PPMI [16] and Fox Insight [15], two landmark studies on Parkinson’s disease. Patients from both datasets presented an age difference between PD and HC; however, differences in age were negligible across datasets. On PPMI, PD participants had an average age of 67.37 ± 9.368 and HC of 56.85 ± 13.90. Similarly, on Fox Insight we observed an average age of 67.68 ± 10.67 for PD participants and 58.06 ± 14.53 for HC.

Questionnaires are often stored as tabular data, which is traditionally analyzed using machine learning methods as they have shown high proficiency at dealing with this type of data [17]. We evaluate Quest2Dx against the most common machine and deep learning methods. These machine learning methods are XGBoost, Random Forest and Support Vector Machine (SVM). More recently, some works have explored using deep learning models such as Multi-layer Perceptrons (MLPs) and Transformers to analyse tabular data. However, some studies [18] have suggested that, while very powerful, these methods might be an overkill for tabular data, requiring more resources and yielding non-statistically significant improvements. However, these studies have been limited to non-medical datasets. On the other hand, transformers have shown to be very effective for PD diagnosis using a diverse set of modalities [19, 20]. Quest2Dx, a transformer model with pre-trained language encoders, outperformed all competing benchmarks, showed strong transferability across datasets, and strong interpretability of the most important questions to identify PD patients.

### 2.1 Screening PD patients from health questionnaires

#### 2.1.1 Data and experimental design

We used data from two landmark datasets for PD, PPMI-online and Fox Insight, which have 40,012 (PD: 27,802, HC: 12,210) and 54,571 (PD: 38,514, HC:16,057) subjects, respectively. Both datasets contain self-reported measurements collected through surveys. The data cover a wide range of questions, including questions regarding life-style, mental status, mood, fine-motor skills, and family history.

Missing entries due to partial completion of questionnaires is a common problem, and health questionnaires are no exception. There are two types of missing entries in the dataset: one occurs when sub-questionnaires were not available to all participants, resulting in systematic missing data for certain questions. The other type arises when participants chose to skip some questions, leading to incomplete responses within the available questionnaires. This study focused on dealing with the second scenario, which is a more common and practical issue in clinical settings. Questions with more than 50% missing responses were excluded from this study, which are mostly add-on questions from auxiliary studies. After that, 187 questions for PPMI and 378 questions for Fox Insight remained. When using a questionnaire with missing responses, traditionally, the mode or mean of the answers for a particular question can be used to impute categorical and continuous data correspondingly. Quest2Dx uses an alternative approach by using a special learnable representation of the missing entries that allows it to better model them. To evaluate this, all methods were compared across several levels of data completeness per patient, ranging from 50% to 90% in increments of 10%. For example, at 70% completeness, each subject had to have at least 70% complete data to be included in the data set. In other words, to have missing less than 30% of its entries. PPMI and Fox Insight have a considerable number of subjects, with different levels of completeness. PPMI presented a much larger change in the number of subjects available at the levels of minimum data completeness compared to Fox Insight. Fig. 2 shows the distribution of completeness for PD and HC in the two datasets. Similarly, the ratio of PD patients to controls is much lower in PPMI than Fox Insight.

**Fig. 2:**
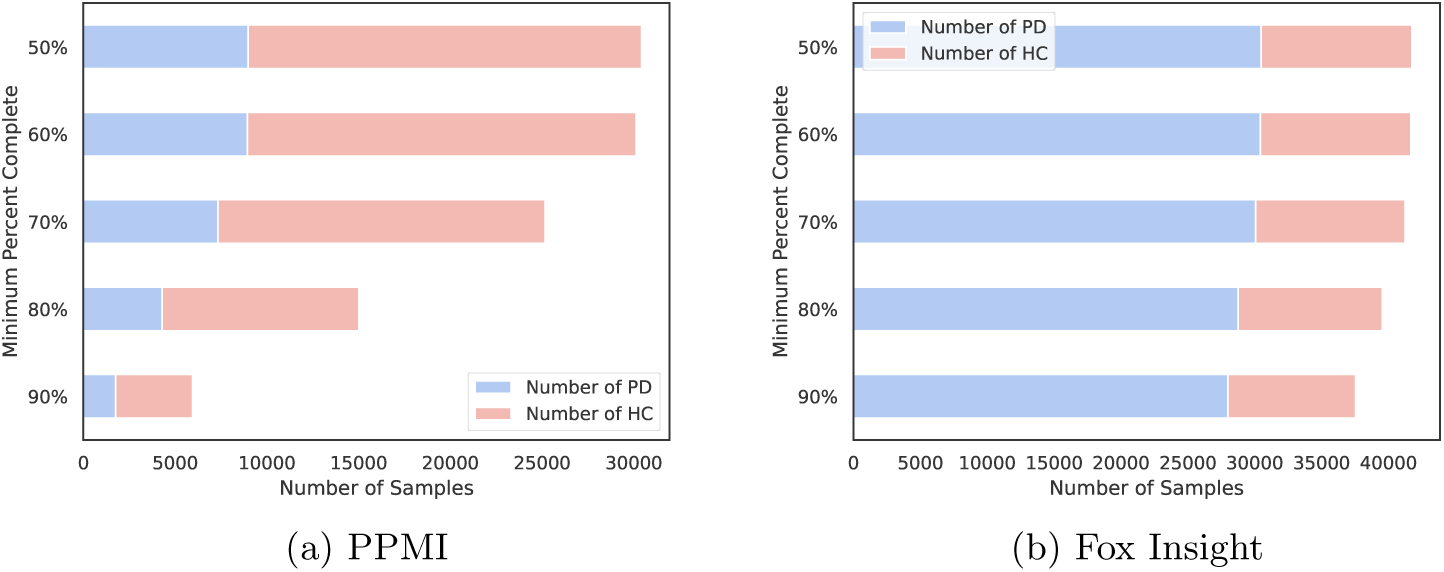
Subject distribution for PPMI and Fox Insight across all levels of minimum data completeness. As the tolerance for less complete data increases, the number of samples available increases reaching a total of 30,432 and 41,755 samples at 50% for PPMI and Fox Insight, respectively. **a.** In PPMI we observe a data imbalance with a larger number of healthy controls than PD patients. **b.** We observed the opposite case on Fox Insight with more PD patients than controls.

All the models were evaluated at five different levels of minimum data completeness using a 5-fold cross-validation. At each one of the data completeness level, for each test fold, we randomly shuffled the training set to train five models for repeated measures. All models were trained and evaluated on the same data splits. This results in a very robust evaluation with a total of 25 experimental runs per model at each data completeness level. We finally reported the average performance.

#### 2.1.2 Quantitative Analysis

Quest2Dx achieved an average AUROC of 0.977 (95% CI: [0.976,0.978]) on PPMI and 0.974 (95% CI: [0.974,0.975]) on Fox Insight for binary classification, surpassing all the comparison benchmarking methods. Comparison of the mean area under the receiver operating characteristic curve (AUROC) for Quest2Dx and benchmarking methods can be found in Fig 3a and Fig 3b. We observe the closest performance to Quest2Dx from the transformer model using mode imputation with an average AUROC of 0.966 on PPMI and 0.971 on Fox Insight. Similarly, the MLP obtained an average AUROC of 0.911 on PPMI and 0.969 on Fox Insight. From the machine learning models, we observed that XGBoost obtained the next highest average AUROC, achieving 0.906 on PPMI and 0.952 on Fox Insight. Random forest performed similarly with an average AUROC of 903 on PPMI and 0.949 on Fox Insight. Finally, the SVM performed the lowest with an average AUROC of 0.888 on PPMI and 0.923 on Fox Insight.

**Fig. 3:**
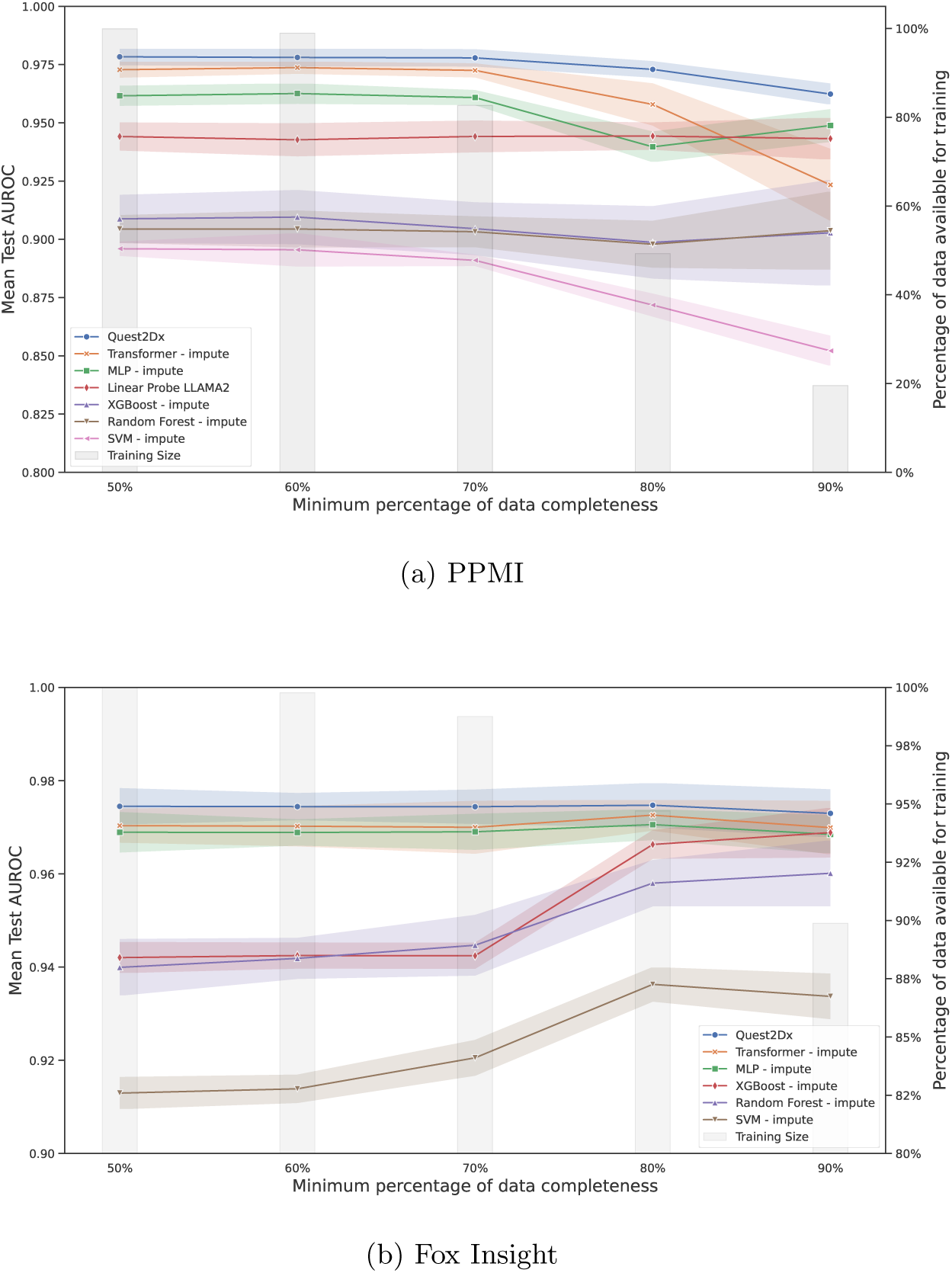
Comparison of Quest2Dx with benchmarking methods. Model performance on PPMI (**a**) and Fox Insight (**b**). The left y-axis shows AUROC scores on test set, while the right y-axis shows the number of samples available for training. Mean AUROC is plotted with lines, while training sample size is shown by the bar plots. The x-axis shows the percentage of minimum samples available at each threshold. The cut off becomes more stringent from left to right and the number of samples for training decreases. Lines represent the mean AUROC across all folds and the shades indicate 95% confidence intervals.

It is important to note that at lower levels of tolerance for missing entries there is less data available. XGBoost and Random Forest maintained stable levels of performance without being greatly affected by the number of missing entries. This is expected as machine learning methods tend to be less affected by the smaller number of samples. SVM had a much lower test accuracy at higher levels of minimum data completeness, namely 90%, 80% and 70%, stabilizing its performance around 60% and 50%. We see similar trends for the MLP and the transformer with mode imputation. We observed that smaller datasets had a considerable impact on the ability of deep learning models to learn on PPMI. Nevertheless, Quest2Dx is able to effectively maintain a reasonably stable performance across all data completeness schedules. The intuition behind this behavior is the infusion of knowledge through embedding the meaning of the questions, allowing the model to learn the representation in a semantic space rather than of memorizing the order of questions. Similarly, the answer content embedding provides prior knowledge regarding the meaning of each value associated with each answer choice. For Fox Insight, we see that all models perform more similar between each other with high AUROCs over 0.900. Compared to PPMI, we see that the number of missing samples has a less drastic drop as the minimum percentage of data completeness increases. Without a sharp drop, the data quality has a considerable role for machine learning models trained on Fox Insight as the performances increase together with the minimum percentage of data completeness.

All models at each level of minimum percentage data completeness were compared to Quest2Dx using a paired t-test. Quest2Dx significantly outperformed all models (*p <* 0.05) at all levels and datasets, except for the transformer model with imputation at 90% completeness on the Fox Insight dataset. We observed clear differences in the models performances between datasets. As seen in 3, the mean test AUROC becomes lower as we approach higher levels of minimum data completeness, whereas in Fox Insight we see increasingly higher mean test AUROCs at higher levels of minimum data completeness. This is due to the data available for training and testing at more strict levels of minimum data completeness. In PPMI we observe a stark decrease in data availability, reaching 20% data availability at 90% minimum data completeness out of the data originally available; whereas in Fox Insight we observe a much smaller decrease. PPMI demonstrates a very realistic scenario where almost complete data is very scarce, and Fox Insight illustrates the ideal case scenario with abundance of data and low levels of imputation. Furthermore, on PPMI, the most important differences are found at high levels of data completeness and low levels of samples available for training, where learning becomes more challenging without the context provided by the BERT [13] question embeddings. By contrast, we observe the smallest difference in performance at high levels of data completeness and negligible decrease in sample size on Fox Insight. These demonstrate the key role played by the knowledge infusion by the BERT embedding and the missing entry learnable representation.

### 2.2 Cross-dataset analysis

A common problem for deep learning models is their limited ability to be applied on unseen datasets from different sources. This could be due to a range of factors such as strong distribution shifts and limited model ability to learn generalizable representations. To evaluate the generalizability of Quest2Dx, we trained it on PPMI and evaluated on Fox Insight, and vice-versa, in a five-fold cross-testing setting. For example, taking PPMI as the training set, PPMI was split in 5 partitions. In each round, 4 partitions were used for training and the remaining one for validation. At each time, Quest2Dx was then evaluated on Fox Insight as the testing set.

We used PPMI as the reference to identify the overlapping questions between the two datasets. The pre-processing of the data includes removing questions that less than 50% of the participants answered. Therefore, questions existing only on PPMI or Fox Insight were not included as part of the training or testing sets. A total of 129 questions overlapped between PPMI and Fox Insight. The overlapping features were identified using the most similar question identified through the cosine similarity and visual inspection to ensure correct matching. Question embeddings similarities are shown in supplementary materials 4.7 As seen in Table 1, as expected there is a performance drop from evaluating Quest2Dx on a independent dataset from the one used for training. Nevertheless, we observe good generalizability from Quest2Dx as it is still able to differentiate PD from HC.

**Table 1:** Comparison of Quest2Dx trained on a dataset and evaluated on a different dataset. Results are shown for evaluation using cross-dataset analysis (e.g. PPMI-Fox Insight and Fox Insight-PPMI). AUROC and 95% CI [lower bound, upper bound] from the 5-fold cross validation is reported for all models.

| Minimum data completeness | Training set size | PPMI<br>→ Fox Insight | Training set size | Fox Insight<br>→ PPMI |
| --- | --- | --- | --- | --- |
| 50% | 24,347 | 0.980 [0.970, 0.991] | 27,060 | 0.945 [0.910, 0.981] |
| 60% | 24,098 | 0.980 [0.973, 0.988] | 19,695 | 0.950 [0.947, 0.952] |
| 70% | 20,082 | 0.973 [0.958, 0.987] | 14,332 | 0.951 [0.939, 0.963] |
| 80% | 15,756 | 0.953 [0.925, 0.981] | 3,747 | 0.973 [0.968, 0.979] |
| 90% | 2,998 | 0.715 [0.696, 0.810] | 3,583 | 0.943 [0.927, 0.959] |

Quest2Dx demonstrated strong generalizability across datasets, achieving an average AUROC of 0.920 (95% CI: [0.741, 1.115]) when trained on PPMI and evaluated on Fox Insight, and 0.952 (95% CI: [0.925, 0.980]) when trained on Fox Insight and tested on PPMI. While these cross-dataset results show a slight decrease compared to the within-dataset evaluations, the performance remains consistent, highlighting the model’s robustness in transferring between datasets.

When trained on PPMI and evaluated on Fox Insight, we observed a similar trend to the within-dataset performance, where AUROCs were lower at 80% and 90% minimum data completeness due to the smaller training set size. However, as more data was available for training and validation on the cross-dataset experiments, Quest2Dx’s performance improved. The model maintained high AUROC scores across different levels of data completeness, with a noticeable drop at 90% completeness, likely due to insufficient data to generalize well. Conversely, when trained on Fox Insight, Quest2Dx maintained consistently strong performance across all levels of data completeness. Similar to the within-dataset evaluation, performance improved with higher levels of data completeness.

### 2.3 Comparison against LLMs on generalization

In recent years, large language models, such as GPT [21], and open source LLAMA [22], have disrupted the natural language processing field as these foundation models allow to process data from multiple sources and for multiple tasks without retraining or finetuning. These models have enabled a wide a range of analysis across multiple fields, from finance to medicine [23]. Similarly, they have been proposed to analyze tabular data showing promising results [24]. The key challenge for their use in tabular data analysis is how to transform the data from its raw form to a suitable input interpretable by an LLM. For example, Chen et al. [25] built a template to better match the flow of a natural sentence and filled it out for each sample. The templates then were used for fact verification, which was implemented as a binary classification problem. One of their findings was the relevance of using templates compared to simple concatenation of the table cells, showing stronger performance.

In this paper, we conducted experiments to evaluate the performance of LLM-based PD patient screening compared to Quest2Dx and machine learning benchmarks. LLAMA2 (7 billion parameter version) was used as the encoder for all subjects to obtain a single vector representation of each. A linear probe was applied on top of the patient representation and trained to classify PD vs controls. As shown in Fig 3a, the LLM-based approached performs comparatively to other deep learning approaches; however, Quest2Dx still has the best performance. As mentioned above, one of the key advantages of LLMs is their ability to encode data from multiple sources. Similarly, as it takes a template-based input, it allows for a flexible number of questions, and missing data. Therefore, we evaluated the generalizability of using a linear probe on LLAMA2. The linear probe was trained on PPMI and evaluated on Fox Insight and vice-versa. Our results showed poor generalizability with random predictions when trained on Fox Insight and evaluated on PPMI 0.568 ± 0.041 and slightly better than random when trained on PPMI and evaluated on Fox Insight, achieving an AUROC of 0.568 *±* 0.071.

### 2.4 Identifying the key questions to screen PD

A key challenge for deep learning methods to be adopted in clinical settings is the lack of interpretability of the model predictions. One of the advantages of transformer-based models is their intrinsic ability to learn the relationship between features (in our case, questions), and the major contributing features to the classification. Through this method, we calculated the contributions of each question-answer pair to the classification and identified the top 10 questions leading to the diagnosis.

Fig. 4 shows that the most important features identified by our model are associated with co-occurring conditions and cognitive decline. For example, difficulty with everyday tasks, such as eating or getting out of bed, may suggest the degeneration of dopamine-producing neurons, which is a hallmark for PD development. Similarly, exposure to solvents and degreasers has been linked to an increased risk of PD [26], as toxins can elevate oxidative stress, contributing to neurodegeneration. While solvent exposure alone is not indicative of PD, it becomes relevant when considered alongside other early signs. This underscores the importance of evaluating multiple factors together.

**Fig. 4:**
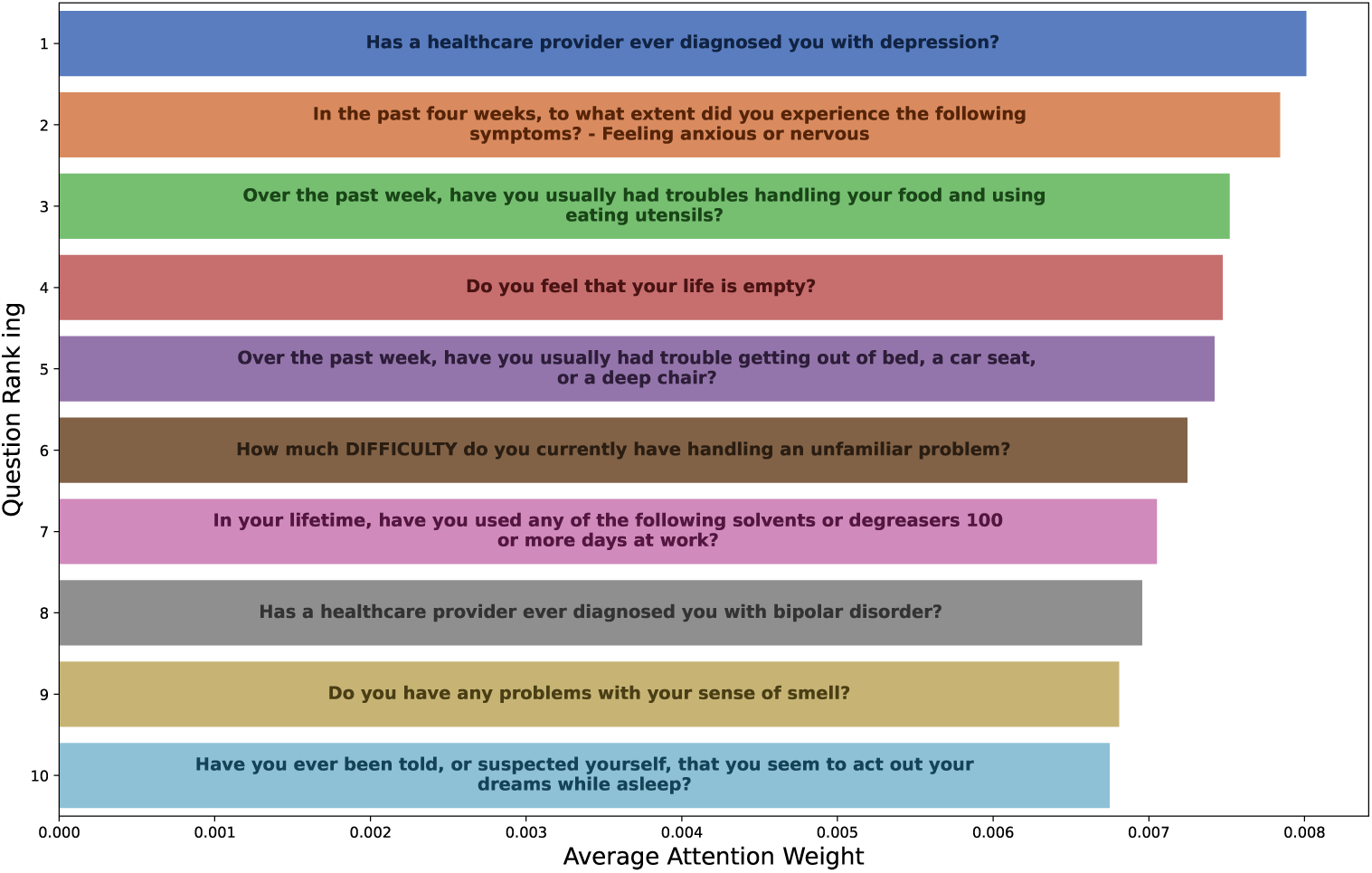
Top 10 questions identified by Quest2Dx on the PPMI dataset by average attention weights. The x-axis shows the average attention weight assigned by Quest2Dx for each question across patients. The higher the value is, the more relevant the feature is towards classifying patients. Y-axis shows the rank with the most important question at the top and the 10*^th^* most important at the bottom.

REM Sleep Behavior Disorder (RBD), where patients act out vivid dreams, is a strong predictor of PD [6]. Thus, being told or suspecting that one acts out their dreams could indicate degeneration in brain stem areas that regulate REM sleep. Anxiety is another common non-motor symptom in PD, often appearing before motor symptoms [6, 27]. Changes in serotonin and dopamine levels contribute to anxiety in PD patients, who are also at increased risk for mood disorders like bipolar disorder due to overlapping pathways involved in dopamine regulation [28].

Feeling tense or stressed is another frequent non-motor symptom in PD and can often exacerbate other symptoms. PD patients often have a heightened stress response due to dysfunction in the hypothalamic-pituitary-adrenal (HPA) axis, leading to elevated cortisol levels [29], which can accelerate neurodegeneration and create a vicious cycle of stress and worsening symptoms.

It is worth noting that using the answer to each one of these questions independently is not sufficient to detect the onset of PD. It is very challenging to screen PD patients based on a single question and its answer. However, the combination of certain answers can be indicative of disease onset. In our work, by analyzing all the exposures, life style changes and early symptoms in conjunction, Quest2Dx is able to screen PD patients effectively.

### 2.5 Effectiveness of questions in classifying PD

While Quest2Dx, and even the benchmarking methods obtain high classification accuracy, they rely on a large number of questions, 187 for PPMI and 378 for Fox Insight. This is a significant limiting factor for the addition of these questions into health questionnaires in clinics. Therefore, we set out to identify a reduced set of questions that still can achieve high classification accuracy. We identified the top 10, 20 and 30 questions used by Quest2Dx on PPMI following the methods described in section 2.4, and created three datasets containing the subsets of questions. All three subset datasets have the same number of samples as the original dataset, since the only modification is the number of questions. Model performance was evaluated using the same 5-fold cross-validations with five repeated measures as introduced in section 2.1.1.

As shown in Fig. 5, while there is a decrease in performance, Quest2Dx is still able to classify PD patients using only 10, 20 and 30 questions. When using only 10 questions, which is about a 5% of the total questions in PPMI, Quest2Dx has a performance drop of about 10%. Using only 20 questions renders reasonable results with AUROC scores over 0.900. Using the top 30 questions still presented a performance degradation; however, it still achieved a high AUROC. We specifically see that at 50% minimum percentage of data completeness the AUROC of using only 30 questions is close to 0.950.

**Fig. 5:**
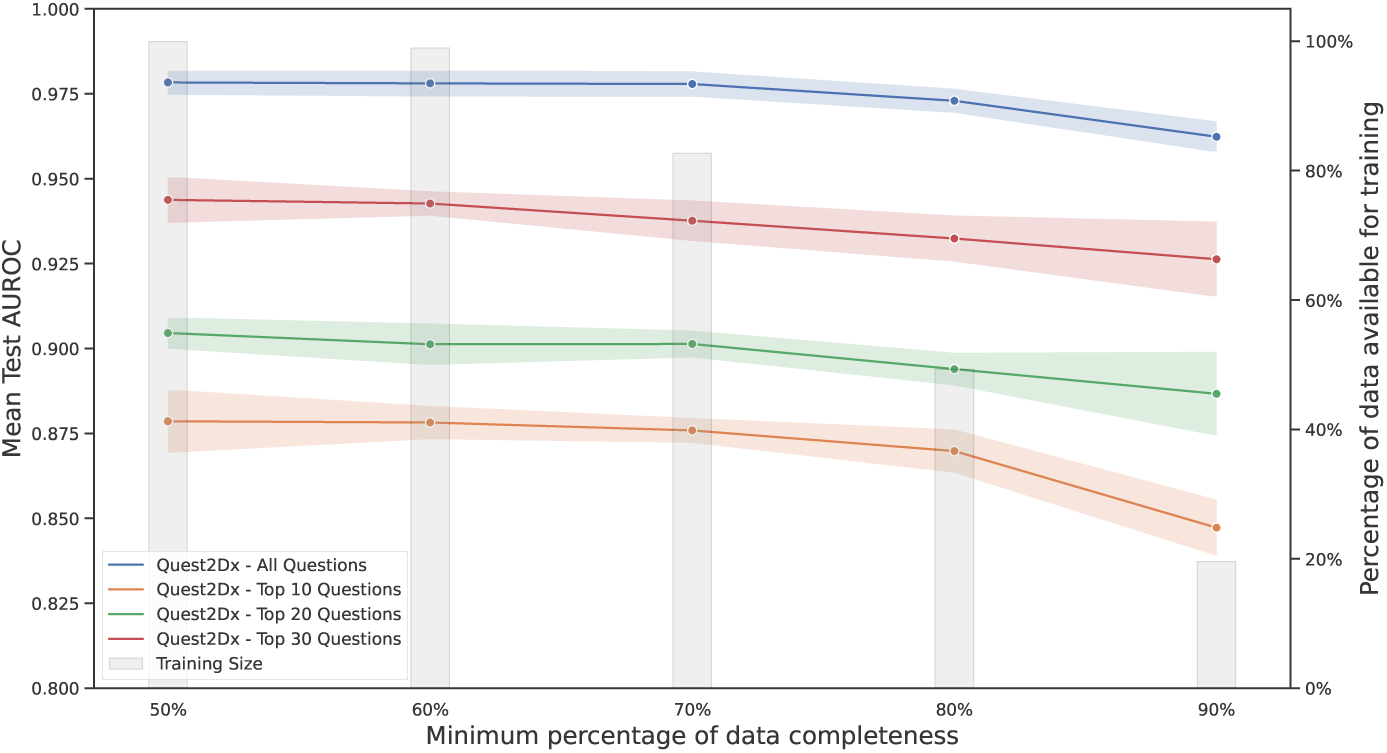
Quest2Dx: Performance comparison of PD classification using all questions vs three subsets. The left y-axis shows AUROC scores on the test set, illustrated by the line plot. While the right y-axis show the percentage of samples available for training, illustrated by the bar plot. The x-axis represents the percentage of minimum samples available at each threshold. The cut off becomes more stringent from left to right and the number of samples for training decreases. Each method is plotted with markers at each matching point on the test AUROC and minimum percentage of data completeness. Lines represent the mean AUROC across all folds and the 95% confidence interval.

## 3 Discussion

Parkinson’s disease screening at early stages is crucial to provide timely care for patients. Early detection of PD can be challenging because the characteristic motor symptoms are not always identifiable at this stage, and other prodromal factors can easily be misinterpreted as other conditions. While a trained specialist can identify these prodromal factors, there is a significant gap between a patient’s initial symptoms and timely referral to a specialist from a primary care setting. Often, patients do not report early-stage symptoms as these can be mistaken for other conditions or ignored. Patients routinely fill out health questionnaires when visiting their primary care physicians, making these questionnaires an crucial potential tool for effective PD screening.

In this study, we demonstrated that both machine learning and deep learning models can classify PD patients from controls using questionnaire data. Deep learning models consistently outperformed machine learning models for both the PPMI and Fox Insight datasets. Specifically, transformer-based models, including our proposed Quest2Dx, outperformed other models, showcasing their capabilities in encoding long-range interactions in the data.

### 3.1 Data Size vs. Data Quality

Our results showed the best performance on Fox Insight in data configurations where the number of missing entries was lowest. This is expected as the models are able to better learn the patterns in data. On PPMI, we see a slightly different trend with lower performance or stricter levels of minimum data completeness (e.g. 90%) where the number of samples available were much smaller (30,432 at 50% v.s. 5,972 90%). This shows a very realistic scenario in which most patients are missing a considerable number of answers. Nevertheless, Quest2Dx was still able to classify the subjects. This finding underscores the importance of data quality and quantity in training effective AI models for PD screening.

### 3.2 Model Performance and Generalizability

Our cross-dataset experiments showed strong performance by Quest2Dx when trained on one dataset and evaluated on the other dataset. We observed stronger performance at higher levels minimum data completeness. This suggests that as the proportion of missing entries grows, Quest2Dx relies more heavily on inferring these gaps from the available data, potentially leading to some degree of overfitting on the training dataset. Alternatively, we used LLAMA2 (7B parameter version) with a linear probe [22] for PD patient screening. Although the linear probe performed comparably well, Quest2Dx consistently outperformed it across all data completeness levels. This suggests that while large language models (LLMs) offer good flexibility in handling data from very different sources, more targeted models like Quest2Dx might be more effective for specific tasks such as PD screening. Improving generalizability is essential for deploying machine learning models in clinical settings.

### 3.3 Key Questions for PD Diagnosis

One of the key advantages of our Quest2Dx model is its ability to identify the most important questions leading to a PD diagnosis. The top questions identified include difficulty in completing daily tasks, exposure to solvents, REM Sleep Behavior Disorder (RBD), anxiety, and mood disorders. These findings align with known comorbidities and prodromal indicators of PD, providing valuable insights for clinical assessments. The identified questions provide valuable insights to what clinicians should pay special attention to during patient evaluations.

### 3.4 Performance of Reduced Questionnaire

We further evaluated the performance of Quest2Dx using subsets of questions identified based on their importance to the performance. The model maintained high classification accuracy even with reduced sets of questions, particularly with the top 30 questions. This demonstrates the potential for implementing a shorter, more practical questionnaire in clinical settings without drastically compromising diagnostic accuracy.

### 3.5 Limitations and Future Directions

Our study has limitations. First, our study only compares PD patients to healthy controls, which simplifies the challenging scenario of distinguishing PD from other neurological disorders that might present overlapping symptoms. While our model was not trained to differentiate PD from conditions like Alzheimer’s disease, the timely referral of these patients to a neurologist would still be beneficial for the patient. Future research should include a broader range of neurodegenerative conditions to enhance the model’s clinical utility by improving its ability to differentiate between similar disorders, ultimately leading to better patient outcomes. Another limitation of Quest2Dx is that it uses self-reported health questionnaire data, which can introduce biases such as recall bias or misunderstanding of questions. This may affect the accuracy of the model’s predictions, as patient-reported outcomes are inherently subjective. While the model performed well on the PPMI and Fox Insight datasets, these datasets may not fully represent the global or more diverse patient populations. Differences in healthcare systems, cultural responses to health questionnaires, or the prevalence of PD symptoms in different populations may limit the generalizability of Quest2Dx.

The Quest2Dx methodology holds great potential for extension to other medical conditions beyond Parkinson’s disease. For instance, it could be adapted to screen for metabolic disorders, such as cardiovascular diseases [30], by analyzing health questionnaire data for early risk factors. Future research could also focus on streamlining the use of reduced questionnaires in clinical settings, particularly for identifying patients in prodromal stages of diseases like PD, thus enhancing early detection and timely intervention. The promising results of Quest2Dx lay a strong foundation for the development of AI-driven screening models based on self-reported data, with the potential to significantly enhance early diagnosis and accessibility across a range of diseases in the future.

## 4 Methods

Our proposed method takes patient reported outcomes as input, learns and classifies the patients as PD or healthy control. Compared to previous approaches, such as machine learning methods, our model learns data representations through a transformer architecture, leverages the power of LLM-learned embeddings and implements a missing entry special token. This special token allows our model to use all the data points available without having to preprocess the data through imputation. Our framework uses BERT [13] to encode the contents and act as knowledge-informed embeddings.

### 4.1 Model architecture

Quest2Dx leverages the feature-tokenizer transformer architecture [18], which has shown strong performances on tabular data modeling. As any transformer it requires converting data into a sequence of tokens with their corresponding embeddings. Both the PPMI and Fox Insight datasets store the patient questionnaire responses in tabular form, as shown in Fig. 6. To prepare this data for the transformer, each subject’s responses were transformed from their categorical (or discrete numerical) format into tokens. These tokens were then mapped to vector representations known as embeddings. In other words, each patient can be thought as a sentence, which is comprised of words. Each “word” of this sentence is an answer whose meaning is encoded in a vector form called embedding. Therefore, each patient is represented as a series of embeddings corresponding to their answers.

**Fig. 6:**
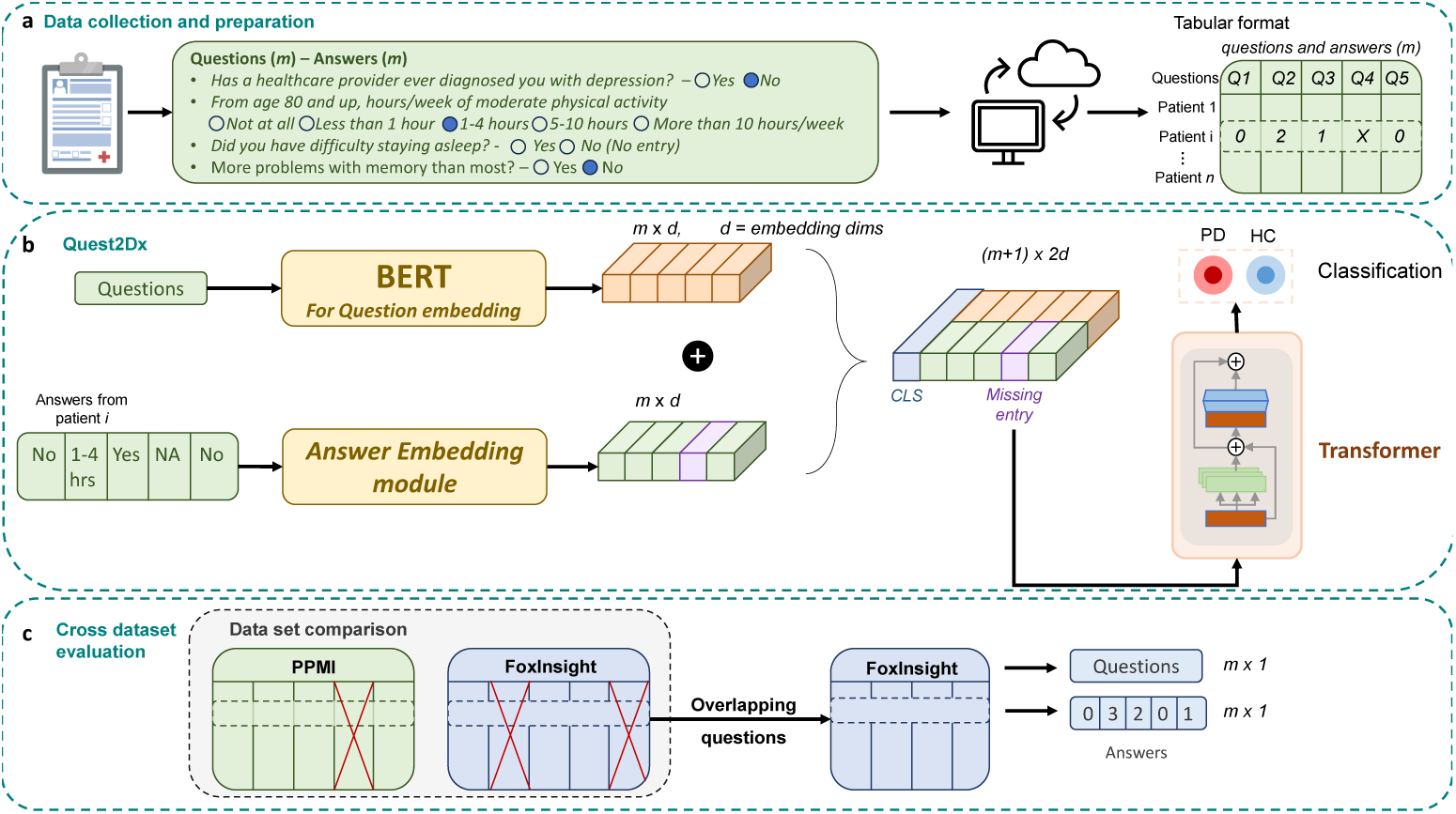
Methods illustration, data, model and evaluation. **a.** Data was collected by PPMI and Fox Insight and stored in tables online. Each table consisted of question IDs as column headers, and participants’ answers coded in numerical form. **b.** Quest2Dx is comprised of 3 main modules, question and answer embedding, transformer model and classification head. Question and answers are embedded separately and the embeddings are then concatenated. The class embedding and missing entry embedding are added to the final patient representation. The transformer models the relationships between answers and learns an overall representation for each subject. Finally, each participant is classified as PD or HC. **c.** The cross dataset evaluation is done by finding the overlapping questions between questionnaires and using the participants data from the question subset to train and validate the model.

To improve the model’s ability to model the patient data, we introduced two special tokens: a class token and a missing entry token. The class token helps capture an overall representation of the patient, enabling more accurate classification. The missing entry token encodes missing data by learning its meaning based on the patterns found in both the patient’s answers and the overall training set. This allows the model to handle incomplete responses without discarding useful information.

To further improve the model’s understanding of the data, we implemented a knowledge-aware encoding mechanism by concatenating BERT embeddings to the answer embeddings. As illustrated in Fig. 6, patient representations are built by concatenating the embeddings of both questions and answers. The process begins with encoding each question from its free-text form into a vector representation (embedding) using BERT. Then, each answer is tokenized and embedded using an embedding module. The question and answer embeddings are concatenated to form a comprehensive question-answer representation for each patient.

These representations are then fed into the transformer model, which learns the latent structure of the data and captures the subject’s health status. Finally, the class token’s embedding is passed through a fully connected layer with a ReLU activation function to classify the subject (e.g., PD or healthy).

### 4.2 Comparison with Alternative Models

For evaluation, Quest2Dx was compared on two aspects, on the model side and the data completeness aspect. On the model side, we compared it against machine learning benchmarks such as Random Forest, Support Vector Machine, XGBoost, and deep learning models, Multi-layer Perceptron and Vanilla Transformer. On the data completeness aspect, we compared against mode-based imputation.

XGBoost is a powerful ensemble learning method that builds a series of decision trees, where each tree corrects the errors of the previous ones, using a technique called gradient boosting. It is highly efficient and performs well on tabular data. The XGBoost used for comparison had 8 estimators, 8 max depth, learning rate of 0.1, and gamma value of 0. It was implemented using the official code [31] and version 2.0.0. Random forest is a well established ensemble method that builds multiple decision trees and combines their predictions. Each tree is trained on a random subset of the data and features, making the model robust and less prone to errors from individual trees. The random forest comparison was implemented using scikit-learn v1.2.0 with hyperparameters of 8 for number of estimators, and a maximum depth of 8. SVM is another well established machine learning method that separates data into classes by finding the optimal boundary that maximizes the margin between different classes. The SVM was implemented using scikit-learn v1.2.0 using a radial basis function kernel and C value of 1. For the deep learning models, we first compared to a simple multilayer perceptron, as it often performs well in a wide variety of tasks. The MLP was implemented using Pytorch and consisted of 2 hidden layers of sizes 128 each. Finally, the vanilla transformer had 4 layers with 4 self-attention heads per layer, and feed-forward layers of dimension of 256.

#### 4.2.1 Model Training and Tuning

Class-weighted cross-entropy was used as the loss and ADAM was used as the optimizer for all deep learning models. Class-weighted cross-entropy was implemented to deal with the imbalance between positive and negative classes. This was particularly helpful for the low sample sizes scenarios. The learning rate for MLP was 0.001, the learning rate for the vanilla transformer was 0.001.

Quest2Dx and all deep learning frameworks were implemented using PyTorch 1.14.0.dev20221026.

Ray-Tune v2.2.0 [32] was used to perform hyperparameter tuning for all models, utilizing the ASHAScheduler, which implements the Async Successive Halving Algorithm. This method allocates a small budget to each configuration and incrementally increases it for the top-performing ones. As resources are allocated, poorly performing configurations are pruned, allowing for efficient hyperparameter optimization across the search space. We used a combination of grid search and random search strategies within Ray-Tune to identify the best set of hyperparameters. For the transformer-based models we tuned the learning rate, drop out, number of layers, attention heads and dimensions of feed-forward layers. For the MLP learning rate and hidden layer dimension sizes were tuned. For the machine learning models we tuned the number of estimators, maximum depth, learning rate, sub sampling rate, gamma, and C regularization terms, and the kernel for the SVM. The best hyperparameters for all models found through Ray-Tune can be found on the supplementary materials.

### 4.3 Cross-Dataset Evaluation

In our cross-dataset evaluation, we opted for static answer embeddings, using pretrained BERT representations for answers rather than dynamic, learnable ones. This approach was selected to provide more flexibility in handling different answer formats across datasets. For example, variations in response categories such as sleep hours (e.g., 1-4, 4-6, 6-8 in one dataset versus 1-4, 4-8, 8+ in another) could still be captured consistently. The special tokens for class and missing entries remained unchanged.

We manually identified overlapping questions between PPMI and Fox Insight and created new subsets for both datasets. A vocabulary was built to map all possible textual answers with their corresponding BERT embeddings. The patient answers were mapped back to their textual form, and using the vocabulary they were mapped into embeddings. As seen in Fig. 6, patient representations were then constructed by combining the class token, the separately encoded BERT-embedded question-answer pairs, and the missing entry embedding when applicable. These representations were fed into the transformer model to classify patients as either PD or healthy controls (HC).

For model training and evaluation, we used 5-fold cross-validation on the training set, while each fold was evaluated on the completely unseen independent test set (e.g. trained on PPMI and evaluated on Fox Insight and vice-versa).

### 4.4 Identifying most relevant questions

Transformer models utilize self-attention to learn a weight matrix that captures how each feature—in this case, each question—relates to one another. The attention mechanism applies a softmax function to the matrix to produce a probability distribution.

The class token is responsible for aggregating all the features to guide patient classification. Consequently, we used the attention values for the class token, which sum to one due to the softmax operation, thereby representing the contribution of each question to the class token. At test time, we extracted the class token values from the last layer of the transformer, summed the attention weights across subjects, and normalized them. Finally, we averaged these values across all cross-validation runs and sorted them to identify the most attended features.

### 4.5 Reduced number of questions

To evaluate the performance of Quest2Dx using a reduced set of questions, three subsets of questions were created to reflect a more streamlined questionnaire suitable for clinical use in screening Parkinson’s disease (PD) patients. These subsets were based on the most important features, as determined in Section 4.4. Specifically, the top 10, 20, and 30 most attended questions were selected to form new datasets, while keeping all other aspects of the original dataset unchanged.

Quest2Dx was then retrained on each of these reduced-question subsets, accounting for the varying levels of missing data present in the entries. The model’s performance was systematically evaluated and compared to its baseline performance, which utilized the full set of questions. The evaluation involved 5 repeated measures of 5-fold cross-validation to ensure robust and reliable results. This approach allowed for an assessment of how reducing the number of questions impacts the model’s predictive accuracy and its potential applicability in real-world clinical settings, where shorter, more focused questionnaires are preferred.

### 4.6 LLM-based classification

First, full patient reports were generated from the tabular data in an effort to recreate the original health questionnaires. To construct the input prompt for the model, each patient’s answers were recoded into their textual form, and the data was structured in a sequential question-answer format. The prompt began with the sentence: “The subject filled a health questionnaire. These are the questions and answers provided by the subject.” This was followed by a list of questions and their corresponding answers, which represented the patient’s responses. The prompt concluded with the question: “Does this patient have Parkinson’s Disease?”. Using this structured input, the LLAMA2 7B (7 billion parameter) model [22] used as text encoder for all subjects to obtain a single vector representation of each one. LLAMA2 7B was obtained from the official MetaAI repository in HuggingFace. Following the recommended approach from MetaAI, as outlined in their HuggingFace documentation, we used the embedding of the last token in the final hidden layer as the classification token. This token serves as the summarized representation of each subject’s data. To enable classification between Parkinson’s disease (PD) patients and healthy controls, a fully connected layer was added on top of the patient representations generated by LLAMA2 7B. This layer was trained to perform binary classification. The training process was conducted using the ADAM optimizer and cross-entropy loss, following the same strategy used for the other evaluated models. This framework allowed for effective training of the classifier, ensuring that the model learned to distinguish between PD patients and control subjects based on the encoded text representations.

### 4.7 Classification Evaluation and Statistical methods

To evaluate the performance of Quest2Dx compared to benchmark models, we used the Area Under the Receiver Operating Characteristic Curve (AUROC) as the primary performance metric. AUROC provides a measure of a model’s ability to distinguish between positive and negative classes, with an AUROC of 1.0 indicating perfect classification and 0.5 indicating random chance. It is defined as the integral of the True Positive Rate (TPR) plotted against the False Positive Rate (FPR) at various thresholds:

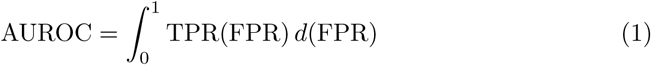

For each model, including Quest2Dx, the AUROC was computed using the roc auc score function from the scikit-learn Python library. The final performance of Quest2Dx was represented as the average AUROC across all experimental runs, which consisted of 5 repeated measures of 5-fold cross-validation. This can be calculated as the mean of the AUROC scores across all folds and repeats

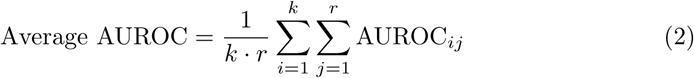

where *k* is the number of folds (5), *r* is the number of repeats (5), and AUROC*_ij_*represents the AUROC score for the *i*-th fold and *j*-th repeat.

To statistically compare the performance of Quest2Dx with benchmark models, we used a paired t-test to assess whether there was a significant difference in the average AUROC between our model and each one of the benchmark models. The null hypothesis *H*_0_ was that there is no difference in the average AUROC between the models, i.e., *µ*_ours_ = *µ*_benchmark_, where *µ* represents the mean AUROC.

The paired t-test was conducted with a significance level *α* = 0.05. The test statistic *t* was calculated as:

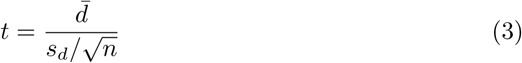

where *d̄* is the mean of the differences between the paired AUROC values, *s_d_* is the standard deviation of these differences, and *n* is the number of paired observations.

We also computed the 95% confidence interval (CI) for the AUROC of each model to estimate the precision of the AUROC values. The confidence intervals were calculated as:

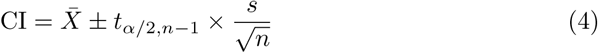

where *X̄* is the sample mean AUROC for a given model, *s* is the sample standard deviation of the AUROC values, and *t_α/_*_2*,n*_*_−_*_1_ is the critical value from the t-distribution with *n −* 1 degrees of freedom.

## Data Availability

The data that support the findings of this study are available from PPMI and Fox Insight but restrictions apply to the availability of these data, which were used under license for the current study, and so are not publicly available. Data are however available from the authors upon reasonable request and with permission of PPMI and Fox Insight.

https://www.ppmi-info.org/

https://foxden.michaeljfox.org/insight/explore/insight.jsp

## Supplementary information

Accompanying supplementary document regarding model hyperparameters and question embedding similarities are included in the submission.

## Acknowledgements

This research was funded in part by the training grant T32AG078123, NSF CAREER award 2046708, and NIH grants U01 AG066833, U01 AG068057, and P30 AG073105. The funder played no role in study design, data collection, analysis and interpretation of data, or the writing of this manuscript.

Data used in the preparation of this article were obtained on November, 13 2024 from the Parkinson’s Progression Markers Initiative (PPMI) database (www. ppmi-info.org/access-data-specimens/download-data), RRID:SCR 006431. For up-to-date information on the study, visit www.ppmi-info.org “PPMI – a public-private partnership – is funded by the Michael J. Fox Foundation for Parkinson’s Research and funding partners, including. PPMI – a public-private partnership – is funded by the Michael J. Fox Foundation for Parkinson’s Research and funding partners, including 4D Pharma, Abbvie, AcureX, Allergan, Amathus Therapeutics, Aligning Science Across Parkinson’s, AskBio, Avid Radiopharmaceuticals, BIAL, BioArctic, Biogen, Biohaven, BioLegend, BlueRock Therapeutics, Bristol-Myers Squibb, Calico Labs, Capsida Biotherapeutics, Celgene, Cerevel Therapeutics, Coave Therapeutics, DaCapo Brainscience, Denali, Edmond J. Safra Foundation, Eli Lilly, Gain Therapeutics, GE HealthCare, Genentech, GSK, Golub Capital, Handl Therapeutics, Insitro, Jazz Pharmaceuticals, Johnson & Johnson Innovative Medicine, Lundbeck, Merck, Meso Scale Discovery, Mission Therapeutics, Neurocrine Biosciences, Neuron23, Neuropore, Pfizer, Piramal, Prevail Therapeutics, Roche, Sanofi, Servier, Sun Pharma Advanced Research Company, Takeda, Teva, UCB, Vanqua Bio, Verily, Voyager v.Therapeutics, the Weston Family Foundation and Yumanity Therapeutics.

The Fox Insight Study (FI) is funded by The Michael J. Fox Foundation for Parkinson’s Research. We would like to thank the Parkinson’s community and 23andMe research participants and employees for making this research possible. Data used in the preparation of this article were obtained from the Fox Insight database (https://foxinsight-info.michaeljfox.org/insight/explore/insight.jsp) on 13/11/2023. For up-to-date information on the study, visit https://foxinsight-info.michaeljfox.org/insight/ explore/insight.jsp

## Declarations

### Ethics approval and consent to participate

Participant consent was obtained by the PPMI and Fox Insight consortiums.

### Consent for publication

All the authors have reviewed the manuscript and agreed to publish the paper once it is accepted.

### Materials availability

Not applicable

### Code availability

The underlying code and training/validation datasets for this study will be made available after publishing.

### Author contribution

Conceptualization, DMR, LS, and PY; Formal analysis, DMR; Funding acquisition, DMR, JH, LS and PY; Investigation, DMR and PY; Methodology, DMR and PY; Experimental design, DMR, JH, LS, and PY; Project administration, PY; Resources, PY; Supervision, PY; Visualization, DMR and PY; Writing—original draft, DMR; Writing—review & editing, JH, LS, and PY.

## Supplementary Information

### Model Hyperparameters

In this section, we provide detailed tables of the hyperparameters used for training our machine learning and deep learning models. The performance of each model can be influenced by the choice of hyperparameters, and careful tuning can be important for achieving optimal results. We employed the ASHAScheduler from Ray-Tune v2.2.0 [32] for hyperparameter optimization, allowing us to efficiently explore a wide range of parameter combinations and identify the best configurations for each model. The tables below summarize the final hyperparameter values selected through this tuning process.

#### Random Forest

**Supplementary Table 1:** Hyperparameters and search space used during the tuning process for the random forest. The table outlines the range or set of possible values for each hyperparameter considered during the Ray- Tune optimization process.

| Hyperparameter | Search space | Value used |
| --- | --- | --- |
| Number of estimators | 2,4,6,8,10 | 10 |
| Maximum depth | 2,4,6,8,10 | 8 |

#### XGBoost

**Supplementary Table 2:**
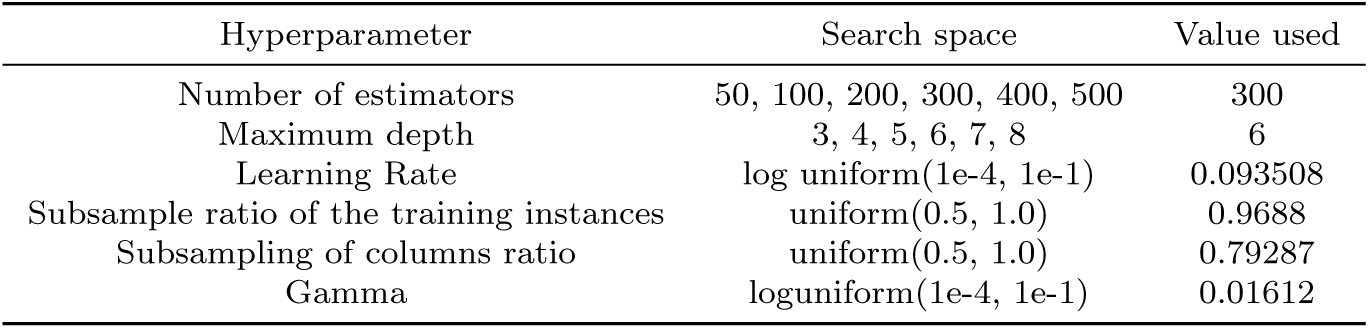
Hyperparameters and search space used during the tuning process for XGBoost. The table outlines the range or set of possible values for each hyperparameter considered during the Ray-Tune optimization process.

#### SVM

**Supplementary Table 3:** Hyperparameters and search space used during the tuning process for the support vector machine (SVM). The table outlines the range or set of possible values for each hyperparameter considered during the Ray-Tune optimization process.

| Hyperparameter | Search space | Value used |
| --- | --- | --- |
| Kernel | RBF, Linear | RBF |
| Regularization parameter C | log uniform(0.1,10) | 0.197273 |

#### MLP

**Supplementary Table 4:** Hyperparameters and search space used during the tuning process for the multi layer preceptron (MLP). The table outlines the range or set of possible values for each hyperparameter considered during the Ray-Tune optimization process.

| Hyperparameter | Search space | Value used |
| --- | --- | --- |
| Learning rate | 0.001,0.0001,0.00001 | 0.0001 |
| Number of units | 32,128,512 | 128 |
| Number of hidden layers | 2,4,6,8 | 2 |

#### Transformer

**Supplementary Table 5:**
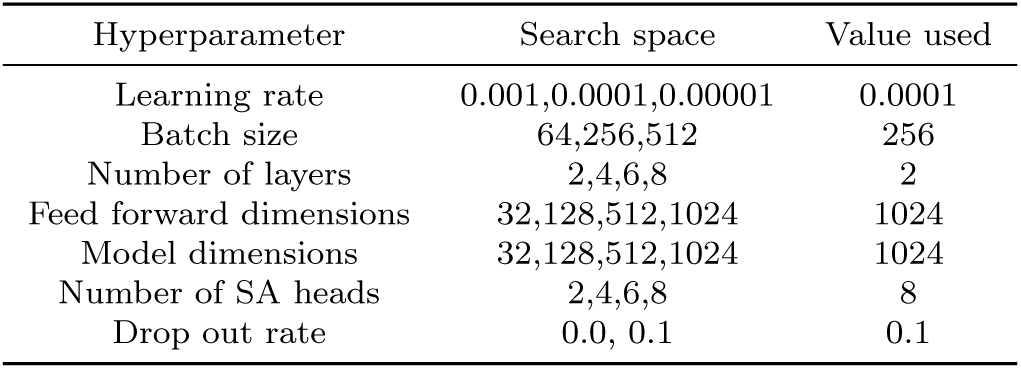
Hyperparameters and search space used during the tuning process for the vanilla feature tokenizer transformer. The table outlines the range or set of possible values for each hyperparameter considered during the Ray-Tune optimization process.

#### Quest2Dx

**Supplementary Table 6:**
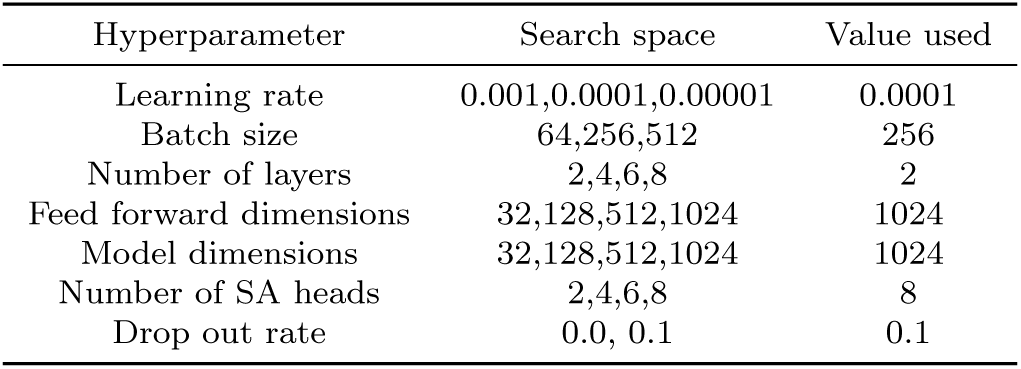
Hyperparameters and search space used during the tuning process for Quest2Dx. The table outlines the range or set of possible values for each hyperparameter considered during the Ray-Tune optimization process.

### Question Similarities

Health questionnaires, particularly those designed for research, can often be extensive and occasionally repetitive in their questions. That is the case for both PPMI Online and Fox Insight datasets, which are composed of several sub-studies, many of which include similar questions. For instance, in Fox Insight, there are questions like *”Have you had anxiety?”* and *”Have you had an anxiety disorder?”* Both inquire about mental health, with the difference being that the first is a general question, while the second specifically refers to a diagnosis.

The question embeddings through BERT allowed not only to enrich the understanding to participants data but to also map similar questions across datasets. In a semantic space questions that refer to similar concepts are closer to each other. For example, first, a few questions could refer to different aspects of sleep, such a quality and how many hours per day in one questionnaire. Second, questions such as as *”Feel that your life is empty?”* on PPMI and *”Do you feel pretty worthless the way you are now?”* on Fox Insight, are slightly different questions but both describe the same mental state of the participant.

To assess the similarity of questions within and between these datasets, we calculated the cosine similarity of the embeddings for every pair of questions. We analyzed the results in two ways: first, by plotting a heatmap of cosine similarities and applying hierarchical clustering to group related questions, and second, by examining the distribution of similarity scores.

#### PPMI

In PPMI, we observed that most questions had a cosine similarity between 0.40 and 0.45. The majority of similarity values were found between 0.2 and 0.6, with the peak around 0.4. Very few questions had high similarity values (i.e. greater than 0.8), with the count dropping significantly as similarity approaches 1.0. This showed a good diversity in the questions contents with only a few ones having a very strong similarity.

**Supplementary Fig. 1:**
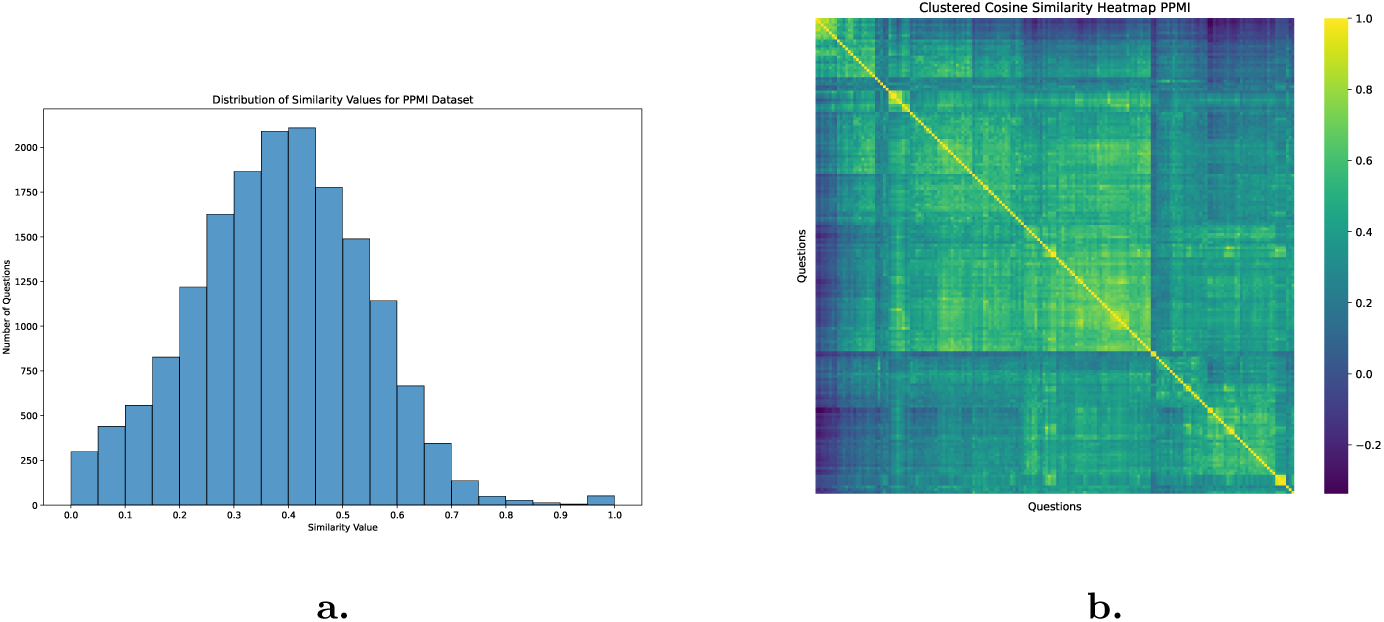
**a.** Cosine similarity score distribution for questions in the PPMI dataset. Histogram shows counts of questions at each similarity bin. Bins are defined at a step of 0.5 from 0.0 to 1.0. **b.** Heat map showing the cosine similarity between questions in PPMI. More similar questions are identified with yellow, while less similar questions are shown in dark blue.

#### Fox Insight

In Fox Insight we observed a distribution more skewed towards lower similarity values indicating more diversity in the content covered by the questions. A majority of the similarity values are clustered around the 0.3 to 0.5 range, with the highest peak slightly above 0.4. Questions with very high similarity values (i.e. above 0.8) are significantly fewer, with those near 1.0 being rare, except for a few ones that were referring to the same question in different sub-questionnaires.

**Supplementary Fig. 2:**
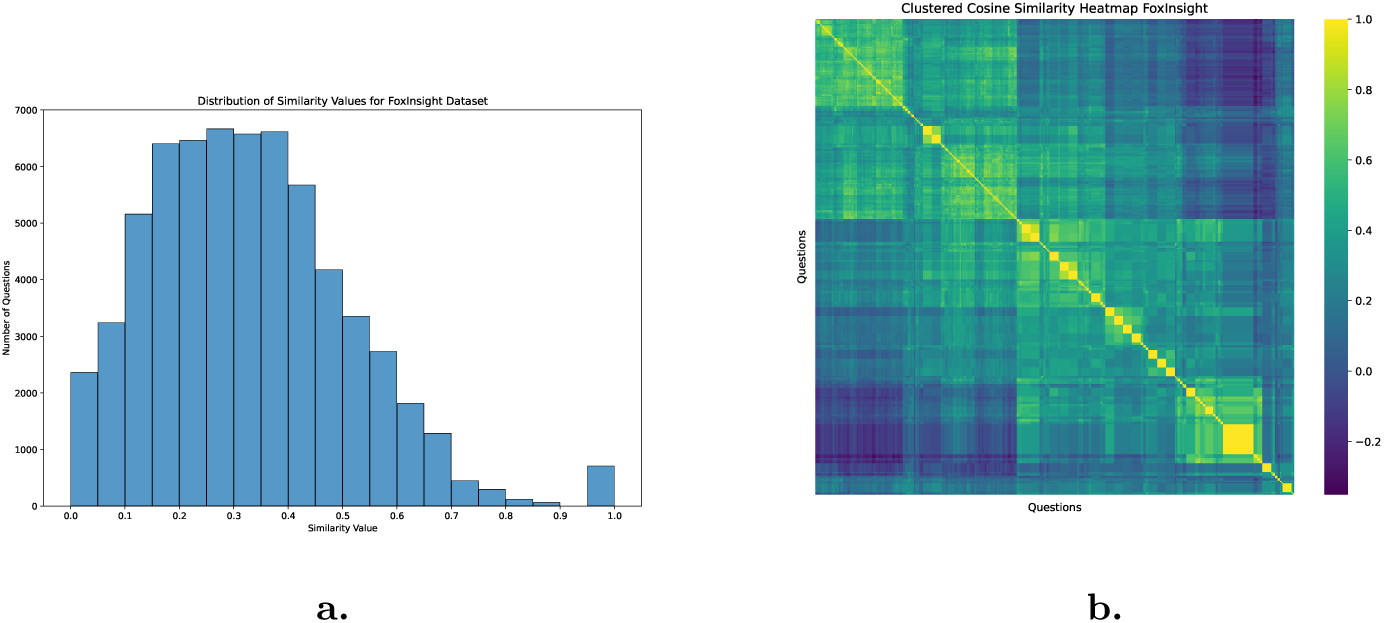
**a.** Cosine similarity score distribution for questions in the Fox Insight dataset. Histogram shows counts of questions at each similarity bin. Bins are defined at a step of 0.5 from 0.0 to 1.0. **b.** Heat map showing the cosine similarity between questions in Fox Insight. More similar questions are identified with yellow, while less similar questions are shown in dark blue.

#### Cross Dataset

When comparing across datasets we observed that 8 features had a cosine similarity of 1.0. These 8 questions had the exact same phrasing on both datasets. Out of the 129 overlapping questions, the same 8 features had a similarity score of above 0.95. 15 questions had at least one similar question on the opposite dataset at 0.85 similarity. 83 and 121 questions had at least one similar question for thresholds 0.65 and 0.50 respectively. We observed that most questions had at least one similar question.

Across datasets we observed some similarities with even a few questions being identically worded. Most of the questions have similarity values between 0.2 and 0.6, with a peak around 0.4, indicating that the majority of cross-dataset questions have moderate similarity. There are few questions with similarity values close to 1.0, showing that identical questions are rare between the two datasets. Nevertheless, we see high abundance of moderately similar questions with the distribution being roughly bell-shaped. Suggesting that while there are some distinct questions across datasets; most of them share similar content.

**Supplementary Fig. 3:**
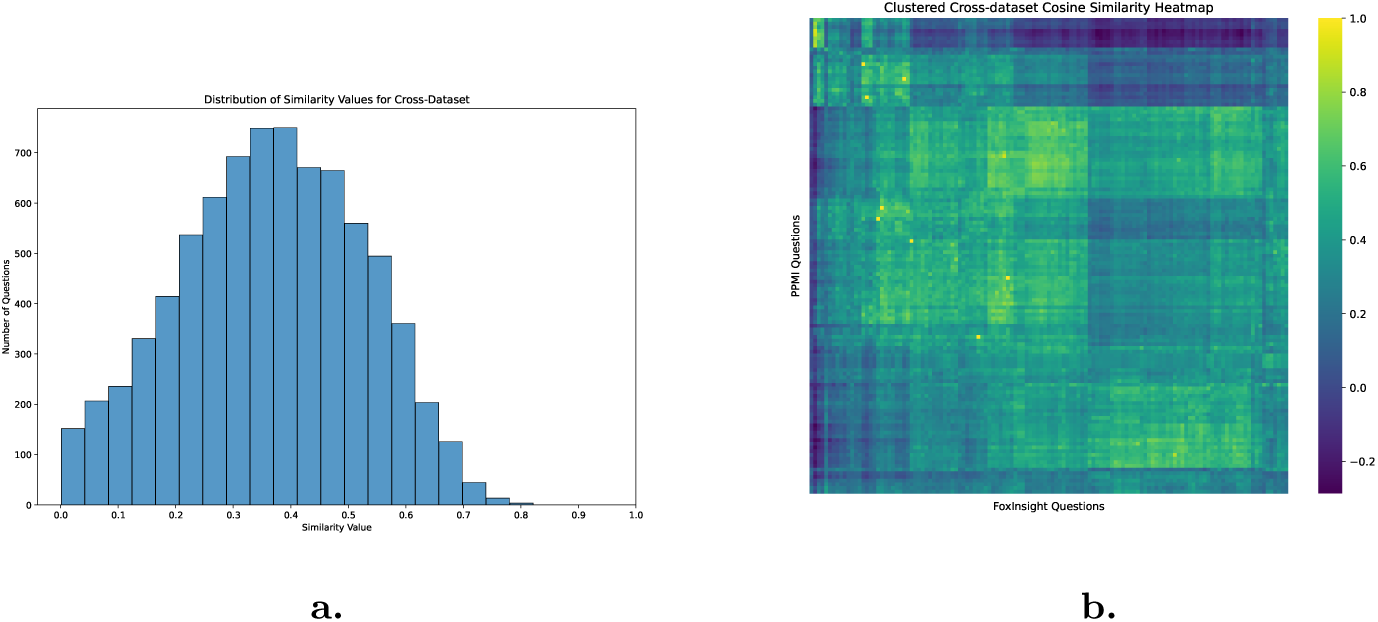
**a.** Cosine similarity score distribution for cross-dataset questions. Histogram shows counts of questions at each similarity bin. Bins are defined at a step of 0.5 from 0.0 to 1.0. **b.** Heat map showing the cosine similarity between questions in PPMI and Fox Insight. More similar questions are identified with yellow, while less similar questions are shown in dark blue.

Overall, the results show that a few groups of highly similar questions exist within each dataset. As expected, we observed a balanced distribution of similarity across PPMI and Fox Insight, with some clusters of related questions. These findings confirm our hypothesis: while there are some in-dataset and cross-dataset questions with overlapping meanings, there are still enough distinct questions to identify patterns that differentiate participants. Additionally, the overlap of information between similar questions contributes to accurate PD prediction, even with a reduced set of questions.

## Question List

### PPMI

1. What is your usual number of bowel movements per day?
2. How often do you typically use laxatives to help you move your bowels?
3. In your lifetime, have you ever regularly drunk hot or iced caffeinated black tea, that is, at least once per WEEK for 6 months or longer?
4. Do you currently regularly drink caffeinated coffee?
5. During the time that you regularly drank caffeinated black tea, on average, how many cups per week did you drink?
6. During the time that you regularly drank caffeinated coffee, on average, how many cups per week did you drink?
7. At what age did you last regularly drink caffeinated coffee?
8. In your lifetime, have you ever regularly drunk caffeinated diet soda, that is, at least once per WEEK for 6 months or longer?
9. In your lifetime, have you ever regularly consumed caffeinated energy drinks or products, that is, at least once per WEEK for 6 months or longer?
10. In your lifetime, have you ever regularly drunk caffeinated green tea, that is, at least once per WEEK for 6 months or longer?
11. Do you currently regularly drink caffeinated regular soda?
12. During the time that you regularly drank caffeinated regular soda, on average, how many cups per week did you drink?
13. In your lifetime, have you ever regularly drunk caffeinated, regular soda, that is, at least once per WEEK for 6 months or longer?
14. At what age did you last regularly drink caffeinated regular soda?
15. In your lifetime, have you used carbon tetrachloride 100 days or more at work
16. In your lifetime, have you used Avgas, jet fuel 100 days or more at work
17. In your lifetime, have you used kerosene 100 days or more at work
18. In your lifetime, have you used methyl ethyl ketone 100 days or more at work
19. In your lifetime, have you used methylene chloride 100 days or more at work
20. In your lifetime, have you used mineral sprits, naptha, or paint thinner 100 days or more at work
21. In your lifetime, have you used n-Hexane 100 days or more at work
22. In your lifetime, did not use solvents or degresers 100 days or more at work
23. In your lifetime, have you used Other solvents or degresers not listed 100 days or more at work
24. In your lifetime, have you used PERC (perchloroethylene) 100 days or more at work
25. In lifetime, used Stoddard solvent 100 days or more at work
26. In your lifetime, have you used toulene 100 days or more at work
27. In your lifetime, have you used trichloroethylene (TCE) 100 days or more at work
28. In your lifetime, not sure if used solvents or degreasers 100 days or more at work
29. In your lifetime, have you used Xylene 100 days or more at work
30. In your lifetime, have you welded metal 100 days or more at work?
31. Have you noticed that you are having more problems with thinking, such as difficulty with memory or concentration, that is a change from your normal abilities?
32. Do you have any family history of Parkinson’s Disease or Parkinsonism?
33. Do any of these family members have Parkinson’s disease or parkinsonism? - Biological Father
34. Do any of these family members have Parkinson’s disease or parkinsonism? - Biological Mother
35. Do any of these family members have Parkinson’s disease or parkinsonism? - None of the above have PD or Parkinsonism”
36. Do any of these family members have Parkinson’s disease or parkinsonism? - q. Prefer not to answer”
37. Do any of these family members have Parkinson’s disease or parkinsonism? - Full Brother
38. Do any of these family members have Parkinson’s disease or parkinsonism? - Full Sister
39. Do any of these family members have Parkinson’s disease or parkinsonism? - Child
40. Do any of these family members have Parkinson’s disease or parkinsonism? - Maternal Half Sibling
41. Do any of these family members have Parkinson’s disease or parkinsonism? - Maternal Aunt or Uncle
42. Do any of these family members have Parkinson’s disease or parkinsonism? - Maternal Cousin
43. Do any of these family members have Parkinson’s disease or parkinsonism? - Paternal Half Sibling
44. Do any of these family members have Parkinson’s disease or parkinsonism? - Paternal Aunt or Uncle
45. Do any of these family members have Parkinson’s disease or parkinsonism? - Paternal Cousin
46. Do any of these family members have Parkinson’s disease or parkinsonism? - Maternal Grandfather
47. Do any of these family members have Parkinson’s disease or parkinsonism? - Maternal Grandmother
48. Do any of these family members have Parkinson’s disease or parkinsonism? - Paternal Grandfather
49. Do any of these family members have Parkinson’s disease or parkinsonism? - Paternal Grandmother
50. Are you afraid that something bad is going to happen to you?
51. Do you think it is wonderful to be alive now?
52. Do you think that most people are better off than you are?
53. Do you often get bored?
54. Have you dropped many of your activities and interests?
55. Do you feel that your life is empty?
56. Do you feel full of energy? No
57. Are you in good spirits most of the time?
58. Do you feel happy most of the time?
59. Do you often feel helpless?
60. Do you prefer to stay at home, rather than going out and doing new things?
61. Do you feel that your situation is hopeless?
62. Do you feel you have more problems with memory than most?
63. Are you basically satisfied with your life?
64. Do you feel pretty worthless the way you are now?
65. Have you ever had a head injury or concussion during your life?
66. Has a healthcare provider ever diagnosed you with anxiety?
67. Have you had your appendix removed?
68. Is bipolar disorder still a problem?
69. Has a healthcare provider ever diagnosed you with chronic fatigue?
70. Do you currently have a diagnosis of any of the following conditions from a physician or other healthcare professional? - Alzheimer’s disease
71. Do you currently have a diagnosis of any of the following conditions from a physician or other healthcare professional? - corticobasal syndrome or degeneration disease
72. Do you currently have a diagnosis of any of the following conditions from a physician or other healthcare professional? - dementia or Lewy Bodies
73. Do you currently have a diagnosis of any of the following conditions from a physician or other healthcare professional? - essential tremor disease
74. Do you currently have a diagnosis of any of the following conditions from a physician or other healthcare professional? - multiple system atrophy disease
75. Do you currently have a diagnosis of any of the following conditions from a physician or other healthcare professional? - Not diagnosed with any of the above conditions
76. Do you currently have a diagnosis of any of the following conditions from a physician or other healthcare professional? - Prefer not to answer diagnosis
77. Do you currently have a diagnosis of any of the following conditions from a physician or other healthcare professional? - progressive supranuclear palsy
78. Has a healthcare provider ever diagnosed you with frozen shoulder?
79. Has a healthcare provider ever diagnosed you with gout?
80. Has a healthcare provider ever diagnosed you with hypertension or high blood pressure?
81. Has a healthcare provider ever diagnosed you with hepatitis?
82. Has a healthcare provider ever diagnosed you with inflammatory bowel disease (such as Crohn’s disease or ulcerative colitis)?
83. Has a healthcare provider ever diagnosed you with melanoma?
84. Have you previously had one or both ovaries surgically removed?
85. Has a healthcare provider ever diagnosed you with peptic ulcer disease?
86. Has a healthcare provider ever diagnosed you with seborrheic dermatitis?
87. Have you previously had a vagotomy procedure (a procedure where the vagus nerve is surgically cut)?
88. Has a healthcare provider ever diagnosed you with depression?
89. Has a healthcare provider ever diagnosed you with diabetes?
90. Has a healthcare provider ever diagnosed you with Erectile dysfunction (ED)?
91. Has a healthcare provider ever diagnosed you with REM sleep behavior disorder, also known as RBD?
92. Do you have any problems with your sense of smell?
93. Over the past week, have you usually had problems dressing? For example, are you slow or do you need help with buttoning, using zippers, putting on or taking off your clothes or jewelry?
94. Over the past week, have you usually had troubles handling your food and using eating utensils? For example, do you have trouble handling finger foods or using forks, knives, spoons, chopsticks?
95. Over the past week, on your usual day when walking, do you suddenly stop or freeze as if your feet are stuck to the floor?
96. Over the past week, have you usually had trouble doing your hobbies or other things that you like to do?
97. Over the past week, have people usually had trouble reading your handwriting?
98. Over the past week, have you usually been slow or do you need help with washing, bathing, shaving, brushing teeth, combing your hair, or with other personal hygiene?
99. Over the past week, have you usually had trouble getting out of bed, a car seat, or a deep chair?
100. Over the past week, have you usually had too much saliva during when you are awake or when you sleep?
101. Over the past week, have you had problems with your speech?
102. Over the past week, have you usually had problems swallowing pills or eating meals? Do you need your pills cut or crushed or your meals to be made soft, chopped, or blended to avoid choking?
103. Over the past week, have you usually had shaking or tremor?
104. Over the past week, do you usually have trouble turning over in bed?
105. Over the past week, have you usually had problems with balance and walking?
106. Over the past week have you had constipation troubles that cause you difficulty moving your bowels?
107. Over the past week, have you usually felt fatigued? This feeling is not part of being sleepy or sad.
108. Over the past week, have you felt faint, dizzy, or foggy when you stand up after sitting or lying down?
109. Over the past week, have you had uncomfortable feelings in your body like pain, aches, tingling, or cramps?
110. Over the past week, have you had trouble staying awake during the daytime?
111. Over the past week, have you had trouble going to sleep at night or staying asleep through the night? Consider how rested you felt after waking up in the morning.
112. Over the past week, have you had trouble with urine control? For example, an urgent need to urinate, a need to urinate too often, or urine accidents?
113. Do you know or have you been told that you ever “act out your dreams” while asleep with very active movements of your arms or legs or loud yelling, screaming or other vocalizations?
114. Have you ever injured yourself or your bed partner during sleep?
115. Have you ever been told you have narcolepsy?
116. Have you ever been told you have sleep apnea?
117. Do you generally sleep with a bed partner?
118. Do you walk in your sleep? “
119. In the past four weeks, to what extent did you experience the following symptoms?

- Feeling anxious or nervous”
120. In the past four weeks, to what extent did you experience the following symptoms?

- Excessive worrying about everyday matters
121. In the past four weeks, to what extent did you experience the following symptoms?

- Fear of something bad, or even the worst, happening
122. In the past four weeks, to what extent did you experience the following symptoms?

- Fear of losing control
123. In the past four weeks, to what extent did you experience the following symptoms?

- Fear or avoid public settings
124. In the past four weeks, to what extent did you experience the following symptoms?

- Fear or avoid social situations
125. In the past four weeks, to what extent did you experience the following symptoms?

- Fear or avoid specific objects or situations
126. In the past four weeks, to what extent did you experience the following symptoms?

- Heart palpitations or heart beating fast
127. In the past four weeks, to what extent did you experience the following symptoms?

- Panic or intense fear
128. In the past four weeks, to what extent did you experience the following symptoms?

- Shortness of breath
129. In the past four weeks, to what extent did you experience the following symptoms?

- Feeling tense or stressed
130. In the past four weeks, to what extent did you experience the following symptoms?

- Being unable to relax
131. Did you have muscle cramps in your arms or legs which woke you up whilst sleeping at night?
132. Did you have difficulty falling asleep each night?
133. Did you have difficulty staying asleep?
134. Did you suffer from distressing dreams at night?
135. Did you suffer from distressing hallucinations at night?
136. Did you feel uncomfortable at night because you were unable to turn around in bed or move due to immobility?
137. Did you feel pain in your arms or legs which woke you up whilst sleeping at night?
138. Did you wake early in the morning with painful posturing of arms and legs?
139. Did you get up at night to pass urine?
140. Did you have restlessness of legs or arms at nights causing disruption of sleep?
141. Overall, did you sleep well during the last week?
142. Did you wake up at night due to snoring or difficulties with breathing?
143. Did you feel tired and sleepy after waking in the morning?
144. On waking, did you experience tremor?
145. Was your sleep disturbed due to an urge to move your legs or arms?
146. Do your arms or legs shake?
147. Do you have more difficulty than you once did typing, using a computer mouse, or using a touchscreen on a tablet or mobile phone?
148. Do your feet ever seem to get stuck to the floor?
149. Do people tell you that your face seems less expressive than it once did?
150. Do you move more slowly than other people your age?
151. Is your balance poor?
152. Do you shuffle your feet and/or take tiny steps when you walk?
153. Do you have trouble buttoning buttons?
154. Do you have trouble rising from a chair?
155. Do people tell you that your voice is softer than it once was?
156. Is your handwriting smaller than it once was?
157. How much DIFFICULTY do you currently have counting the correct amount of money when making purchases?
158. How much DIFFICULTY do you currently have discussing a TV show, book, movie, or current events?
159. How much DIFFICULTY do you currently have explaining how to do something involving several steps to another person?
160. How much DIFFICULTY do you currently have doing more than one thing at a time?
161. How much DIFFICULTY do you currently have learning to use new gadgets or machines around the house?
162. How much DIFFICULTY do you currently have reading and following complex instructions (e.g. directions for a new medication)?
163. How much DIFFICULTY do you currently have reading the newspaper or magazine?
164. How much DIFFICULTY do you currently have remembering what day and month it is?
165. How much DIFFICULTY do you currently have remembering a list of 4 or 5 errands without writing it down?
166. How much DIFFICULTY do you currently have remembering new information like phone numbers or simple instructions?
167. How much DIFFICULTY do you currently have keeping track of time (e.g. using a clock)?
168. How much DIFFICULTY do you currently have maintaining or completing a train of thought?
169. How much DIFFICULTY do you currently have understanding your personal financial affairs?
170. How much DIFFICULTY do you currently have handling an unfamiliar problem (e.g. getting the refrigerator fixed)?
171. How much DIFFICULTY do you currently have using a map to tell where to go?
172. Over your lifetime, have you ever had a JOB in which you mixed, applied, or were exposed in some other way to any type of pesticide, including herbicides (kill weeds), fungicides (kill fungus/mold), insecticides (kill insects), or fumigants (gas used to kill fungus/mold or insects)?
173. Acting Out Dreams
174. Have you ever been told, or suspected yourself, that you seem to “act out your dreams” while asleep (for example punching, flailing your arms in the air, making running movements, etc.)?

### FoxInsight

1. Do you currently have a form of heart disease?
2. What kind of heart disease do you have? - Congestive heart failure
3. What kind of heart disease do you have? - Valvular heart disease
4. What kind of heart disease do you have? - Arrhythmia
5. What kind of heart disease do you have? - Coronary heart disease
6. What kind of heart disease do you have? - Atrial fibrillation
7. What kind of heart disease do you have? - Other
8. Do you currently have high blood pressure?
9. Do you currently have lung disease (not cancer)?
10. What kind of lung disease did you have? - Asthma
11. What kind of lung disease did you have? - Emphysema
12. What kind of lung disease did you have? - Chronic obstructive pulmonary disease (COPD)
13. What kind of lung disease did you have? - Pneumonia
14. What kind of lung disease did you have? - Tuberculosis
15. Do you currently have diabetes?
16. Do you currently have gastric disturbances (not cancer)?
17. What type of gastric disturbances did you have? - Acid reflux (GERD)
18. Have you had gastric disturbances (not cancer)? - Gastritis
19. Have you had gastric disturbances (not cancer)? - Hiatal hernia
20. Have you had gastric disturbances (not cancer)? - Ulcer
21. Do you currently have kidney disease (not cancer)?
22. Do you currently have kidney disease (not cancer)? - Renal failure
23. Do you currently have kidney disease (not cancer)? - Cysts
24. Do you currently have kidney disease (not cancer)? - Kidney stones
25. Do you currently have liver disease (not cancer)?
26. Do you currently have liver disease (not cancer)? - Cirrhosis
27. Do you currently have liver disease (not cancer)? - Chronic viral hepatitis (Hepatitis C or hep C)
28. Do you currently have liver disease (not cancer)? - Hepatitis A
29. Do you currently have liver disease (not cancer)? - Hepatitis B
30. Do you currently have a blood disease (not cancer)?
31. Do you currently have a blood disease (not cancer)? - Anemia
32. Do you currently have a blood disease (not cancer)? - Thalassemia
33. Do you currently have a blood disease (not cancer)? - Sickle cell disease
34. Do you currently have cancer?
35. What type of cancer do you currently have - Bladder
36. What type of cancer do you currently have - Breast
37. What type of cancer do you currently have - Colon
38. What type of cancer do you currently have - Leukemia
39. What type of cancer do you currently have - Liver (Hepatic cancer)
40. What type of cancer do you currently have - Lung
41. What type of cancer do you currently have - Lymphoma
42. What type of cancer do you currently have - Melanoma
43. What type of cancer do you currently have - Prostate
44. What type of cancer do you currently have - Thyroid
45. What type of cancer do you currently have - Skin (non-melanoma)
46. What type of cancer do you currently have - Uterine
47. Do you currently have depression?
48. Do you currently have arthritis?
49. What type of arthritis did you have? - Osteoarthritis/degenerative arthritis
50. What type of arthritis did you have? - Rheumatoid arthritis
51. Have you had back pain lasting longer than a week?
52. Have you had anxiety?
53. Because of the Parkinson’s Disease, how much DIFFICULTY do you currently have reading the newspaper or magazine?
54. How much DIFFICULTY do you currently have keeping track of time (e.g. using a clock)?
55. How much DIFFICULTY do you currently have counting the correct amount of money when making purchases?
56. How much DIFFICULTY do you currently have reading and following complex instructions (e.g. directions for a new medication)?
57. How much DIFFICULTY do you currently have handling an unfamiliar problem (e.g. getting the refrigerator fixed)?
58. How much DIFFICULTY do you currently have explaining how to do something involving several steps to another person?
59. How much DIFFICULTY do you currently have remembering a list of 4 or 5 errands without writing it down?
60. How much DIFFICULTY do you currently have using a map to tell where to go?
61. How much DIFFICULTY do you currently have remembering new information like phone numbers or simple instructions?
62. How much DIFFICULTY do you currently have doing more than one thing at a time?
63. How much DIFFICULTY do you currently have learning to use new gadgets or machines around the house?
64. How much DIFFICULTY do you currently have understanding your personal financial affairs?
65. How much DIFFICULTY do you currently have maintaining or completing a train of thought?
66. How much DIFFICULTY do you currently have discussing a TV show, book, movie, or current events?
67. How much DIFFICULTY do you currently have remembering what day and month it is?
68. Please indicate your preferences in the use of hands in the following object: Spoon
69. Have you ever had a form of heart disease?
70. Have you ever had a heart attack?
71. Have you ever had high blood pressure?
72. Have you ever had lung disease (not cancer)?
73. Have you ever had diabetes?
74. Have you had gastric disturbances (not cancer)?
75. Have you ever had kidney disease (not cancer)?
76. Have you ever had liver disease (not cancer)?
77. Have you ever had a blood disease (not cancer)?
78. Have you ever had cancer?
79. Kidney (Renal cancer)
80. Have you had depression?
81. Have you had arthritis?
82. Have you ever had a stroke (including TIA or transient ischemic attack)?
83. Have you had a traumatic brain injury (TBI)?
84. Have you had any surgeries that required anesthesia?
85. What type(s) of surgery have you had? - Cardiac surgery
86. What type(s) of surgery have you had? - Orthopaedic surgery
87. What type(s) of surgery have you had? - Gastrointestinal surgery
88. What type(s) of surgery have you had? - Cranial or brain surgery
89. What type(s) of surgery have you had? - Tumor removal
90. What type(s) of surgery have you had? - Pulmonary (lung) surgery
91. What type(s) of surgery have you had? - ENT surgery
92. What type(s) of surgery have you had? - Eye surgery
93. What type(s) of surgery have you had? - Reproductive surgery
94. What type(s) of surgery have you had? - Cosmetic surgery
95. Are you basically satisfied with your life?
96. Have you dropped many of your activities and interests?
97. Do you feel that your life is empty?
98. Do you often get bored?
99. Are you in good spirits most of the time?
100. Are you afraid that something bad is going to happen to you?
101. Do you feel happy most of the time?
102. Do you often feel helpless?
103. Do you prefer to stay at home, rather than going out and doing new things?
104. Do you feel you have more problems with memory than most people?
105. Do you think it is wonderful to be alive?
106. Do you feel pretty worthless the way you are now?
107. Do you feel full of energy?
108. Do you feel that your situation is hopeless?
109. Do you think that most people are better off than you?
110. Who is filling out this questionnaire?
111. Speech: Over the past week, have you had problems with your speech?
112. Saliva and Drooling: Over the past week, have you usually had too much saliva during when you are awake or when you sleep?
113. Chewing and Swallowing: Over the past week, have you usually had problems swallowing pills or eating meals? Do you need your pills cut or crushed or your meals to be made soft, chopped or blended to avoid choking?
114. Eating Tasks: Over the past week, have you usually had troubles handling your food and using eating utensil? For example, do you have trouble handling finger foods or using forks, knifes, spoons, chopsticks?
115. Dressing: Over the past week, have you usually had problems dressing? For example, are you slow or do you need help with buttoning, using zippers, putting on or taking off your clothes or jewelry?
116. Hygiene: Over the past week, have you usually beenslow or do you need help with washing, bathing, shaving, brushing teeth, combing your hair or with other personal hygiene?
117. Handwriting: Over the past week, have people usually had trouble reading your handwriting?
118. Doing hobbies or other activities: Over the past week, have you usually had trouble doing your hobbies or other things that you like to do?
119. Turning in Bed: Over the past week, have you usually have trouble turning over in bed?
120. Temor: Over the past week, have you usually had shaking or tremor?
121. Getting out of bed, a care, or a deep chair: Over the past week, have you usually had trouble getting out of a bed, a car seat, or a deep chair?
122. Walking and Balance: Over the past week, have you usually had problems with balance and walking?
123. Freezing: Over the past week, on your usual day when walking, do you suddently stop or freeze as if your feet are stuck to the floor?
124. Do you have a family history of Parkinson’s disease?
125. Which family members have/had Parkinson’s disease? - Mother
126. Which family members have/had Parkinson’s disease? - Father
127. Which family members have/had Parkinson’s disease? - Child
128. Which family members have/had Parkinson’s disease? - Grandchild
129. Which family members have/had Parkinson’s disease? - Great-Grandchild
130. Which family members have/had Parkinson’s disease? - Sibling
131. Which family members have/had Parkinson’s disease? - Half-Sibling
132. Which family members have/had Parkinson’s disease? - Maternal Grandmother
133. Which family members have/had Parkinson’s disease? - Maternal Grandfather
134. Which family members have/had Parkinson’s disease? - Maternal Aunt
135. Which family members have/had Parkinson’s disease? - Maternal Uncle
136. Which family members have/had Parkinson’s disease? - Maternal Cousin
137. Which family members have/had Parkinson’s disease? - Maternal Niece/Nephew
138. Which family members have/had Parkinson’s disease? - Paternal Grandmother
139. Which family members have/had Parkinson’s disease? - Paternal Grandfather
140. Which family members have/had Parkinson’s disease? - Paternal Aunt
141. Which family members have/had Parkinson’s disease? - Paternal Uncle
142. Which family members have/had Parkinson’s disease? - Paternal Cousin
143. Which family members have/had Parkinson’s disease? - Paternal Niece/Nephew
144. Do you have a family history of Alzheimer’s disease, dementia or memory loss?
145. Do you have a family history of Amyotrophic Lateral Sclerosis (ALS)?
146. Do you have a family history of dystonia (painful, prolonged muscle contractions that cause involuntary repetitive twisting and sustained muscle contractions)?
147. Do you have a family history of bi-polar disorder or schizophrenia?
148. Do you have a family history of depression?
149. Do you have a family history of anxiety?
150. Have you experienced dribbling of saliva during the daytime in the last month?
151. Have you experienced loss or change in your ability to taste or smell in the last month?
152. Have you experienced difficulty swallowing food or drink or problems with choking in the last month?
153. Have you experienced vomiting or feelings of sickness (nausea) in the last month?
154. Have you experienced constipation (less than three bowel movements a week) or having to strain to pass a stool in the last month?
155. Have you experienced bowel (fecal) incontinence in the last month?
156. Have you experienced feeling that your bowel emptying is incomplete after having been to the toilet in the last month?
157. Have you experienced a sense of urgency to pass urine that makes you rush to the toilet in the last month?
158. Have you experienced getting up regularly at night to pass urine in the last month?
159. Have you experienced unexplained pains (not due to known conditions such as arthritis) in the last month?
160. Have you experienced unexplained change in weight (not due to change in diet) in the last month?
161. Have you experienced problems remembering things that have happened recently or forgetting to do things in the last month?
162. Have you experienced loss of interest in what is happening around you or in doing things in the last month?
163. Have you experienced seeing or hearing things that you know or are told are not there in the last month?
164. Have you experienced difficulty concentrating or staying focused in the last month?
165. Have you experienced feeling sad, ‘low’ or ‘blue’ in the last month?
166. Have you experienced feeling anxious, frightened or panicky in the last month?
167. Have you experienced feeling less interested in sex or more interested in sex in the last month?
168. Have you experienced finding it difficult to have sex when you try in the last month?
169. Have you experienced feeling light-headed, dizzy or weak standing from sitting or lying in the last month?
170. Have you experienced falling in the last month?
171. Have you experienced finding it difficult to stay awake during activities such as working, driving or eating in the last month?
172. Have you experienced difficulty getting to sleep at night or staying asleep at night in the last month?
173. Have you experienced intense, vivid or frightening dreams in the last month?
174. Have you experienced talking or moving about in your sleep, as if you are ‘acting out’ a dream in the last month?
175. Have you experienced unpleasant sensations in your legs at night or while resting, and a feeling that you need to move in the last month?
176. Have you experienced swelling of the legs in the last month?
177. Have you experienced excessive sweating in the last month?
178. Have you experienced double vision in the last month?
179. Have you experienced believing things are happening to you that other people say are not in the last month?
180. Leisure time activity: Over the past 7 days, how often did you particpate in activities such as reading, watching TV or doing handcrafts?
181. On average, how many hours per day did you enage in sitting activities?
182. Walking activities: Over the past 7 days, how often did you take a walk outside your home or yard for any reason? For example, for fun or exercise, walking to work, walking the dog etc.?
183. On average, how many hours per day did you spend walking?
184. Light sport and recreational activities: Over the past 7 days, how often did you engage in light sport or recreational activities such as bowling, gold with a cart, shuffleboard, fishing from a boat or pier or other similar activities?
185. Moderate sport and recreational activities: Over the past 7 days, how often did you engage in moderate sport recreational activities such as doubles tennis, ball-room dancing, hunting, ice skating, gold without a care, softball or other similar activities?
186. Strenuous sport and recreational activities: Over the past 7 days, how often did you engage in strenuous sport activities such as jogging, swimming, cycling, singles tennis, aerobic dance, skiiing (downhill or cross country) or other similar activities?
187. Muscle strength: Over the past 7 days, how often did you do exercise specifically to increase muscle strength and endurance, such as lifting weights or push-ups etc.?
188. Household activity: During the past 7 days, have you done any light housework, such as dusting or washing dishes?
189. Household activity: During the past 7 days, have you done any heavy housework or chores such as vacuuming, scrubbing floors, washing windows, or carrying wood?
190. Household activity: During the past 7 days, did you engage in home repairs like painting, wallpapering, electrical work etc.?
191. Household activity: During the past 7 days, did you engage in lawn work or yard care, including snow or leaf removal, wood chopping, etc.?
192. Household activity: During the past 7 days, did you engage in outdoor gardening?
193. Household activity: During the past 7 days, did you engage in caring for another person, such as children, dependent, spouse, or another adult?
194. Work-Related Activity: During the past 7 days, did you work for pay or as a volunteer? “
195. Mobility: Please tick the one box that best describes your health today describes your “
196. Self-Care: Please tick the one box that best describes your health toda
197. Usual activities (e.g. work, study, housework, family or leisure activities): Please tick the one box that best describes your health today
198. Pain/discomfort: Please tick the one box that best describes your health today
199. Anxiety/depression: Please tick the one box that best describes your health today
200. We would like to know how good or bad your health is TODAY. Think about your health on a scale numbered from 0 to 100. Now, please write the number that best represents your health TODAY in the box below
201. Have you ever been told, or suspected yourself, that you seem to ‘act out your dreams’ while asleep (for example, punching, flailing your arms in the air, making running movements, etc.)?

### Overlap

| PPMI | FoxInsight |
| --- | --- |
| Over your lifetime, have you ever had a JOB in which you mixed, applied, or were exposed in some other way to any type of pesticide, including herbicides (kill weeds), fungicides (kill fungus/-mold), insecticides (kill insects), or fumigants (gas used to kill fungus/mold or insects)? | Over your lifetime, have you ever had a JOB in which you mixed, applied, or were exposed in some other way to any type of pesticide, including herbicides (kill weeds), fungicides (kill fungus/-mold), insecticides (kill insects), rodenticides (kill rats/mice), or fumigants (gas |
| Who is completing this questionnaire? | Who is filling out this questionnaire? |
| In your lifetime, have you welded metal 100 days or more at work? | In your lifetime, have you welded, brazed, or flame cut metal 100 or more days at work or at home? |
| In your lifetime, have you used Other solvents or degreasers not listed 100 days or more at work | What specific solvents or degreasers did/do you use? - Other Solvent |
| In your lifetime, have you used Xylene 100 days or more at work | What specific solvents or degreasers did/do you use? - Toluol/toluene/xylol/xylene |
| In your lifetime, have you used trichloroethylene (TCE) 100 days or more at work | What specific solvents or degreasers did/do you use? - Trichloroethane |
| In your lifetime, have you used PERC (perchloroethylene) 100 days or more at work | What specific solvents or degreasers did/do you use? - Trichloroethylene or PERC (perchloroethylene) |
| In your lifetime, have you used n-Hexane 100 days or more at work | What specific solvents or degreasers did/do you use? - n-Hexane |
| In your lifetime, have you used mineral sprits, naptha, or paint thinner 100 days or more at work | What specific solvents or degreasers did/do you use? - Mineral spirits, naptha, paint thinner |
| In your lifetime, have you used methyl ethyl ketone 100 days or more at work | What specific solvents or degreasers did/do you use? - Methyl ethyl ketone (MEK) |
| In your lifetime, have you used methylene chloride 100 days or more at work | In your lifetime, have you used solvents or degreasers 100 or more days at work or at home? (Such as carbon tetrachloride, chloroform, methylene chloride, trichloroethane, trichloroethylene or PERC (perchloroethylene), acids, alkalis, alcohols, avgas, jet fuel, freon, methyl ethyl ketone (MEK), kerosene, mineral spirits, naptha, paint thinner, n-Hexane, Stoddard solvent, toluol/toluene/xylol/xulene, turpentine) |
| In your lifetime, have you used kerosene 100 days or more at work | What specific solvents or degreasers did/do you use? |
| In your lifetime, have you used Avgas, jet fuel 100 days or more at work | What specific solvents or degreasers did/do you use? - Avgas, jet fue |
| Has a healthcare provider ever diagnosed you with hypertension or high blood pressure? | Do you currently have high blood pressure? |
| Have you ever had a head injury or concussion during your life? | Have you ever had a head injury or concussion? |
| During the time that you regularly drank caffeinated regular soda, on average, how many cups per week did you drink? | During the time you were regularly drinking caffeinated soda, on average, about how many cups per week did you drink? |
| At what age did you last regularly drink caffeinated regular soda? | Age started regularly drinking caffeinated soda |
| In your lifetime, have you ever regularly drunk caffeinated green tea, that is, at least once per WEEK for 6 months or longer? | In your lifetime, have you ever regularly drunk caffeinated green tea, that is, at least once per week for 6 months or longer? |
| In your lifetime, have you ever regularly drunk hot or iced caffeinated black tea, that is, at least once per WEEK for 6 months or longer? | In your lifetime, have you ever regularly drunk hot or iced caffeinated black tea, that is, at least once per week for 6 months or longer? (black tea includes most types of non-herbal tea, such as Lipton, Earl Grey and others) |
| During the time that you regularly drank caffeinated black tea, on average, how many cups per week did you drink? | During the time you were regularly drinking caffeinated coffee, on average, about how many cups per week did you drink? |
| At what age did you last regularly drink caffeinated coffee? | Age started regularly drinking caffeinated coffee |
| During the time that you regularly drank caffeinated coffee, on average, how many cups per week did you drink? | In your lifetime, have you ever regularly drunk caffeinated coffee, that is, at least once per week for 6 months or longer? |
| Has a healthcare provider ever diagnosed you with hepatitis? | Do you currently have liver disease (not cancer)? - Chronic viral hepatitis (Hepatitis C or hep C) |
| Have you had your appendix removed? | Have you had an appendix removal? |
| Have you previously had one or both ovaries surgically removed? | Have you had an ovary surgery? |
| Over the past week, on your usual day when walking, do you suddenly stop or freeze as if your feet are stuck to the floor? | Freezing: Over the past week, on your usual day when walking, do you suddenly stop or freeze as if your feet are stuck to the floor? |
| Has a healthcare provider ever diagnosed you with melanoma? | What type of cancer do you currently have - Melanoma |
| Is bipolar disorder still a problem? | Has a Physician diagnosed you with this condition? - Bipolar disorder |
| Over the past week, have you usually had problems with balance and walking? | Walking and Balance: Over the past week, have you usually had problems with balance and walking? |
| Over the past week, have you usually had trouble getting out of bed, a car seat, or a deep chair? | Getting out of bed, a care, or a deep chair: Over the past week, have you usually had trouble getting out of a bed, a car seat, or a deep chair? |
| Over the past week, have you usually had shaking or tremor? | Temor: Over the past week, have you usually had shaking or tremor? |
| Over the past week, do you usually have trouble turning over in bed? | Turning in Bed: Over the past week, have you usually have trouble turning over in bed? |
| How much DIFFICULTY do you currently have remembering what day and month it is? | How much DIFFICULTY do you currently have remembering what day and month it is? |
| Over the past week, have you usually had trouble doing your hobbies or other things that you like to do? | Doing hobbies or other activities: Over the past week, have you usually had trouble doing your hobbies or other things that you like to do? |
| Did you wake up at night due to snoring or difficulties with breathing? | Please check all of the symptoms that you experience during OFF periods - Trouble breathing |
| Has a healthcare provider ever diagnosed you with anxiety? | Has a Physician diagnosed you with this condition? - Anxiety diagnosis |
| Do you think that most people are better off than you are? | Do you think that most people are better off than you? |
| Over the past week, have people usually had trouble reading your handwriting? | Handwriting: Over the past week, have people usually had trouble reading your handwriting? |
| How much DIFFICULTY do you currently have discussing a TV show, book, movie, or current events? | How much DIFFICULTY do you currently have discussing a TV show, book, movie, or current events? |
| Do you feel that your situation is hopeless? | Do you feel that your situation is hopeless? |
| Over the past week, have you usually been slow or do you need help with washing, bathing, shaving, brushing teeth, combing your hair, or with other personal hygiene? | Hygiene: Over the past week, have you usually been slow or do you need help with washing, bathing, shaving, brushing teeth, combing your hair or with other personal hygiene? |
| How much DIFFICULTY do you currently have maintaining or completing a train of thought? | How much DIFFICULTY do you currently have maintaining or completing a train of thought? |
| Over the past week, have you usually had problems dressing? For example, are you slow or do you need help with buttoning, using zippers, putting on or taking off your clothes or jewelry? | Dressing: Over the past week, have you usually had problems dressing? For example, are you slow or do you need help with buttoning, using zippers, putting on or taking off your clothes or jewelry? |
| How much DIFFICULTY do you currently have understanding your personal financial affairs? | How much DIFFICULTY do you currently have understanding your personal financial affairs? |
| Do you move more slowly than other people your age? | Do you move more slowly than other people your age? |
| Do you feel full of energy? No | Do you feel full of energy? |
| In the past four weeks, to what extent did you experience the following symptoms? - Fear or avoid specific objects or situations | In the past four weeks, to what extent did you fear or avoid the following situations? - Specific objects or situations (such as flying, heights, spiders or other animals, needles, or blood) |
| How much DIFFICULTY do you currently have learning to use new gadgets or machines around the house? | How much DIFFICULTY do you currently have learning to use new gadgets or machines around the house? |
| Over the past week, have you usually had troubles handling your food and using eating utensils? For example, do you have trouble handling finger foods or using forks, knives, spoons, chopsticks? | Eating Tasks: Over the past week, have you usually had troubles handling your food and using eating utensil? For example, do you have trouble handling finger foods or using forks, knives, spoons, chopsticks? |
| Do you feel pretty worthless the way you are now? | Do you feel pretty worthless the way you are now? |
| How much DIFFICULTY do you currently have doing more than one thing at a time? | How much DIFFICULTY do you currently have doing more than one thing at a time? |
| Do you shuffle your feet and/or take tiny steps when you walk? | Do you shuffle your feet and/or take tiny steps when you walk? |
| Do you have trouble buttoning buttons? | Do you have trouble buttoning buttons? |
| Do you think it is wonderful to be alive now? | Do you think it is wonderful to be alive? |
| Over the past week, have you usually had problems swallowing pills or eating meals? Do you need your pills cut or crushed or your meals to be made soft, chopped, or blended to avoid choking? | Chewing and Swallowing: Over the past week, have you usually had problems swallowing pills or eating meals? Do you need your pills cut or crushed or your meals to be made soft, chopped or blended to avoid choking? |
| In the past four weeks, to what extent did you experience the following symptoms? - Fear or avoid public settings | In the past four weeks, to what extent did you fear or avoid the following situations? - Public settings (situations from which it may be difficult or embarrassing to escape, such as queues or lines, crowds, bridges, or public transportation) |
| How much DIFFICULTY do you currently have remembering new information like phone numbers or simple instructions? | How much DIFFICULTY do you currently have remembering new information like phone numbers or simple instructions? |
| Over the past week, have you usually had too much saliva during when you are awake or when you sleep? | Saliva and Drooling: Over the past week, have you usually had too much saliva during when you are awake or when you sleep? |
| In the past four weeks, to what extent did you experience the following symptoms? - Fear or avoid social situations | In the past four weeks, to what extent did you fear or avoid the following situations? - Social situations (where one may be observed, or evaluated by others, such as speaking in public, or talking to unknown people) |
| Do you feel you have more problems with memory than most? | Do you feel you have more problems with memory than most people? |
| Do your arms or legs shake? | Do your arms or legs shake? |
| Do people tell you that your face seems less expressive than it once did? | Do people tell you that your face seems less expressive than it once did? |
| Do you prefer to stay at home, rather than going out and doing new things? | Do you prefer to stay at home, rather than going out and doing new things? |
| Over the past week, have you had problems with your speech? | Speech: Over the past week, have you had problems with your speech? |
| Did you get up at night to pass urine? | Have you experienced getting up regularly at night to pass urine in the last month? |
| How much DIFFICULTY do you currently have using a map to tell where to go? | How much DIFFICULTY do you currently have using a map to tell where to go? |
| In the past four weeks, to what extent did you experience the following symptoms? - Fear of losing control | In the past four weeks, did you experience episodes of the following symptoms? - Fear of losing control |
| Over the past week, have you usually felt fatigued? This feeling is not part of being sleepy or sad. | Over the past week, have you usually felt fatigued? This feeling is not part of being sleepy or sad |
| In the past four weeks, to what extent did you experience the following symptoms? - Heart palpitations or heart beating fast | In the past four weeks, did you experience episodes of the following symptoms? - Heart palpitations or heart beating fast (not related to physical effort or activity) |
| How much DIFFICULTY do you currently have remembering a list of 4 or 5 errands without writing it down? | How much DIFFICULTY do you currently have remembering a list of 4 or 5 errands without writing it down? |
| Do you often feel helpless? | Do you often feel helpless? |
| Has a healthcare provider ever diagnosed you with depression? | Have you had depression? |
| Do your feet ever seem to get stuck to the floor? | Do your feet ever seem to get stuck to the floor? |
| How much DIFFICULTY do you currently have explaining how to do something involving several steps to another person? | How much DIFFICULTY do you currently have explaining how to do something involving several steps to another person? |
| In the past four weeks, to what extent did you experience the following symptoms? - Shortness of breath | In the past four weeks, did you experience episodes of the following symptoms? - Shortness of breath |
| Over the past week, have you felt faint, dizzy, or foggy when you stand up after sitting or lying down? | In the past four weeks, did you experience episodes of the following symptoms? - Lightheadedness with standing/orthostatic hypotension/fainting |
| Do you currently have a diagnosis of any of the following conditions from a physician or other healthcare professional? - Alzheimer's disease | Is there a medical or psychiatric condition that would preclude participation? - Alzheimer's disease |
| Did you suffer from distressing dreams at night? | Have you experienced intense, vivid or frightening dreams in the last month? |
| Do you feel happy most of the time? | Do you feel happy most of the time? |
| Is your balance poor? | Is your balance poor? |
| How much DIFFICULTY do you currently have handling an unfamiliar problem (e.g. getting the refrigerator fixed)? | How much DIFFICULTY do you currently have handling an unfamiliar problem (e.g. getting the refrigerator fixed)? |
| In the past four weeks, to what extent did you experience the following symptoms? - Panic or intense fear | In the past four weeks, did you experience episodes of the following symptoms? - Panic or intense fear |
| Are you afraid that something bad is going to happen to you? | Are you afraid that something bad is going to happen to you? |
| Do you currently have a diagnosis of any of the following conditions from a physician or other healthcare professional? - corticobasal syndrome or degeneration disease | Is there a medical or psychiatric condition that would preclude participation? - Cortical Basal Ganglionic Degeneration |
| Over the past week have you had constipation troubles that cause you difficulty moving your bowels? | Have you experienced constipation (less than three bowel movements a week) or having to strain to pass a stool in the last month? |
| In the past four weeks, to what extent did you experience the following symptoms? -Fear of something bad, or even the worst, happening | Please score to what extent you experienced the following symptoms in the past four weeks - Fear of something bad, or even the worst, happening |
| How much DIFFICULTY do you currently have reading and following complex instructions (e.g. directions for a new medication)? | How much DIFFICULTY do you currently have reading and following complex instructions (e.g. directions for a new medication)? |
| Do people tell you that your voice is softer than it once was? | Do people tell you that your voice is softer than it once was? |
| Are you in good spirits most of the time? | Are you in good spirits most of the time? |
| Do you currently have a diagnosis of any of the following conditions from a physician or other healthcare professional? - multiple system atrophy disease | Is there a medical or psychiatric condition that would preclude participation? - Multiple system atrophy or MSA |
| How much DIFFICULTY do you currently have counting the correct amount of money when making purchases? | How much DIFFICULTY do you currently have counting the correct amount of money when making purchases? |
| In the past four weeks, to what extent did you experience the following symptoms? - Excessive worrying about everyday matters | Please score to what extent you experienced the following symptoms in the past four week - Excessive worrying about everyday matters |
| Do you often get bored? | Do you often get bored? |
| Do you currently have a diagnosis of any of the following conditions from a physician or other healthcare professional? - progressive supranuclear palsy | Is there a medical or psychiatric condition that would preclude participation? - Progressive supranuclear palsy or PSP |
| Is your handwriting smaller than it once was? | Is your handwriting smaller than it once was? |
| Did you have difficulty staying asleep? | Have you experienced difficulty getting to sleep at night or staying asleep at night in the last month? |
| How much DIFFICULTY do you currently have keeping track of time (e.g. using a clock)? | How much DIFFICULTY do you currently have keeping track of time (e.g. using a clock)? |
| In the past four weeks, to what extent did you experience the following symptoms? - Being unable to relax | Please score to what extent you experienced the following symptoms in the past four weeks - Being unable to relax |
| Over the past week, have you had trouble staying awake during the daytime? | Please score to what extent you experienced the following symptoms in the past four weeks - Daytime sleepiness |
| Do you currently have a diagnosis of any of the following conditions from a physician or other healthcare professional? - dementia or Lewy Bodies | Is there a medical or psychiatric condition that would preclude participation? - Dementia with Lewy Bodies or DLB |
| Do you feel that your life is empty? | Do you feel that your life is empty? |
| Have you ever been told, or suspected yourself, that you seem to "act out your dreams" while asleep (for example punching, flailing your arms in the air, making running movements, etc.)? | Have you ever been told, or suspected yourself, that you seem to 'act out your dreams' while asleep (for example, punching, flailing your arms in the air, making running movements, etc.)? |
| In the past four weeks, to what extent did you experience the following symptoms? - Feeling tense or stressed | Please score to what extent you experienced the following symptoms in the past four weeks - Feeling tense or stressed |
| How much DIFFICULTY do you currently have reading the newspaper or magazine? | Because of the Parkinson's Disease, how much DIFFICULTY do you currently have reading the newspaper or magazine? |
| Did you have difficulty falling asleep each night? | In the past month, have you had trouble falling asleep when you went to bed at night? |
| Do you have any problems with your sense of smell? | Have you experienced loss or change in your ability to taste or smell in the last month? |
| Have you dropped many of your activities and interests? | Have you dropped many of your activities and interests? |
| Do you currently have a diagnosis of any of the following conditions from a physician or other healthcare professional? - essential tremor disease | Is there a medical or psychiatric condition that would preclude participation? - Essential tremor |
| Has a healthcare provider ever diagnosed you with diabetes? | Have you had diabetes? |
| Do you have any family history of Parkinson's Disease or Parkinsonism? | Do you have a family history of Parkinson's disease? |
| In the past four weeks, to what extent did you experience the following symptoms? - 1. Feeling anxious or nervous | Please score to what extent you experienced the following symptoms in the past four weeks - Feeling anxious or nervous |
| Are you basically satisfied with your life? | Are you basically satisfied with your life? |
| Have you ever been told you have sleep apnea? | Have you had sleep apnea? |
| Has a healthcare provider ever diagnosed you with seborrheic dermatitis? | Have you had seborrheic dermatitis? |
| Has a healthcare provider ever diagnosed you with inflammatory bowel disease (such as Crohn's disease or ulcerative colitis)? | Have you had irritable bowel syndrome (IBS)? |
| Has a healthcare provider ever diagnosed you with peptic ulcer disease? | Have you had gastric disturbances (not cancer)? - Ulcer |
| Do any of these family members have Parkinson's disease or parkinsonism? - Child | Which family members have/had Parkinson's disease? - Child |
| Do any of these family members have Parkinson's disease or parkinsonism? - Paternal Aunt or Uncle | Which family members have/had Parkinson's disease? - Paternal Aunt |
| Do any of these family members have Parkinson's disease or parkinsonism? - Maternal Cousin | Which family members have/had Parkinson's disease? - Maternal Cousin |
| Do any of these family members have Parkinson's disease or parkinsonism? - Paternal Cousin | Which family members have/had Parkinson's disease? - Paternal Cousin |
| Do any of these family members have Parkinson's disease or parkinsonism? - Maternal Half Sibling | Which family members have/had Parkinson's disease? - Half-Sibling |
| Do any of these family members have Parkinson's disease or parkinsonism? - Maternal Aunt or Uncle | Which family members have/had Parkinson's disease? - Maternal Aunt |
| Do any of these family members have Parkinson's disease or parkinsonism? - Paternal Grandfather | Which family members have/had Parkinson's disease? - Paternal Grandfather |
| Do any of these family members have Parkinson's disease or parkinsonism? - Full Sister | Which family members have/had Parkinson's disease? - Sibling |
| Do any of these family members have Parkinson's disease or parkinsonism? - Paternal Grandmother | Which family members have/had Parkinson's disease? - Paternal Grandmother |
| Do any of these family members have Parkinson's disease or parkinsonism? - Maternal Grandfather | Which family members have/had Parkinson's disease? - Maternal Grandfather |
| Do any of these family members have Parkinson's disease or parkinsonism? - Maternal Grandmother | Which family members have/had Parkinson's disease? - Maternal Grandmother |
| Do any of these family members have Parkinson's disease or parkinsonism? - Biological Father | Which family members have/had Parkinson's disease? - Father |
| Do any of these family members have Parkinson's disease or parkinsonism? - Biological Mother | Which family members have/had Parkinson's disease? - Mother |

